# GRPa-PRS: A risk stratification method to identify genetically-regulated pathways in polygenic diseases

**DOI:** 10.1101/2023.06.19.23291621

**Authors:** Xiaoyang Li, Brisa S. Fernandes, Andi Liu, Jingchun Chen, Xiangning Chen, Zhongming Zhao, Yulin Dai

## Abstract

**Background:** Polygenic risk scores (PRS) are tools used to evaluate an individual’s susceptibility to polygenic diseases based on their genetic profile. A considerable proportion of people carry a high genetic risk but evade the disease. On the other hand, some individuals with a low risk of eventually developing the disease. We hypothesized that unknown counterfactors might be involved in reversing the PRS prediction, which might provide new insights into the pathogenesis, prevention, and early intervention of diseases.

**Methods:** We built a novel computational framework to identify genetically-regulated pathways (GRPas) using PRS-based stratification for each cohort. We curated two AD cohorts with genotyping data; the discovery (disc) and the replication (rep) datasets include 2722 and 2854 individuals, respectively. First, we calculated the optimized PRS model based on the three recent AD GWAS summary statistics for each cohort. Then, we stratified the individuals by their PRS and clinical diagnosis into six biologically meaningful PRS strata, such as AD cases with low/high risk and cognitively normal (CN) with low/high risk. Lastly, we imputed individual genetically-regulated expression (GReX) and identified differential GReX and GRPas between risk strata using gene-set enrichment and variational analyses in two models, with and without *APOE* effects. An orthogonality test was further conducted to verify those GRPas are independent of PRS risk. To verify the generalizability of other polygenic diseases, we further applied a default model of GRPa-PRS for schizophrenia (SCZ).

**Results:** For each stratum, we conducted the same procedures in both the disc and rep datasets for comparison. In AD, we identified several well-known AD-related pathways, including amyloid-beta clearance, tau protein binding, and astrocyte response to oxidative stress. Additionally, we discovered resilience-related GRPs that are orthogonal to AD PRS, such as the calcium signaling pathway and divalent inorganic cation homeostasis. In SCZ, pathways related to mitochondrial function and muscle development were highlighted. Finally, our GRPa-PRS method identified more consistent differential pathways compared to another variant-based pathway PRS method.

**Conclusions:** We developed a framework, GRPa-PRS, to systematically explore the differential GReX and GRPas among individuals stratified by their estimated PRS. The GReX-level comparison among those strata unveiled new insights into the pathways associated with disease risk and resilience. Our framework is extendable to other polygenic complex diseases.

## INTRODUCTION

Polygenic risk score (PRS) estimates an individual’s genetic predisposition to polygenic diseases by aggregating the effects of numerous common genetic variants [1–3]. For instance, Alzheimer’s disease (AD), a progressive neurodegenerative condition primarily affecting the elderly [4], has a genetic architecture composed of common variants [5–8], rare variants [9,10], and copy number variants [11]. The current PRS prediction performance on pathologically confirmed late-onset Alzheimer’s disease (LOAD) cases and controls could reach 0.75–0.84 based on the area under the receiver operating curve (AUROC) [12,13]. Among the common variants, the *APOE4* allele is the strongest risk factor, with individuals with this variant showing higher odds for AD with one *APOE4* allele (odds ratio [OR] 4.6 [95% confidence interval (CI) 4.1–5.2]) or two *APOE4* alleles (OR 25.3 [95%CI 20.4–31.2]) compared with those without the *APOE4* allele [14]. However, *APOE* allele alone does not fully account for LOAD development, prompting ongoing research into additional genetic factors and pathways [15–17]. Schizophrenia (SCZ) is a severe mental disorder marked by disturbances in thought processes, emotions, and behavior, primarily affecting adults. Compared to AD, SCZ is more polygenic with no prominent genetic risk factor like *APOE* to AD [18,19]. Nonetheless, previous research indicates that rare variants play a significant role in SCZ pathogenesis [20]. The SCZ PRS prediction performance could reach 0.71 to 0.74 [21,22]. However, both environmental and genetic factors might influence the prediction of PRS. For instance, individuals with high education attainment had a lower risk of AD and lower cognitive decline, even if they had a high PRS for AD [23]. Recently, Hess et al. [24] and Hou et al. [25] conducted GWAS on high-risk cases vs high-risk controls for SCZ and AD, respectively. Through the analyses, they identified genetic factors associated with resilience as counterfactors orthogonal to PRS liability. Therefore, characterizing these orthogonal genetic factors and related biological functions might provide novel insights into disease pathogenesis and early intervention, including lifestyle modifications and pharmacological interventions [2,26].

To understand the functional enrichment of genetic risks, methods like MAGMA [27] and PRSet [28] aggregate variant effects to the gene level and estimate the gene sets/pathways enrichment of genetic factors. Both MAGMA and PRSet account for the potential inter-gene correlation within the gene-set analysis or inter-variant correlation within each gene set, respectively. Moreover, MAGMA provides both self-contained and competitive gene-set analysis for differential genes derived from case-control comparison. In contrast, PRSet calculates the individual polygenic scores for each gene set of interest in a single-sample manner of gene set analysis [29]. However, genetic variants of complex diseases are context-specific, methodologies like transcriptome-wide association study (TWAS) can incorporate context-specific expression and show better power to unveil novel disease susceptibility loci [30–34], playing an important role in bridging common variants to their genetically-regulated expression (GReX) and providing additional insights not possible with genotyping alone. TWAS-GSEA [35] method was developed for the downstream analysis of the TWAS/FUSION method [36]. It accounts for inter-gene correlations by calculating them from the tissue expression data used to construct the FUSION model and treating these correlations as a random effect in the linear mixed model. However, two potential issues arise: 1) As suggested by MAGMA [27], the inter-gene correlation is corrected for their association with the disease, rather than the inter-gene correlation derived from tissue expression; 2) TWAS-GSEA requires the input FUSION model and is not generalizable to other TWAS methods such as PrediXcan [37] or MultiXcan [38].

In this work, we developed a novel computational framework (GRPa-PRS) to systematically explore differential GReX and GRPa among individuals based on their PRS risk strata, using AD and SCZ as exemplars. Specifically, we aimed to 1) develop new computational methods to identify differential GRPa from AD or SCZ strata comparison; 2) disentangle the GRPa without the *APOE* factor in AD; 3) identify the GRPa that is orthogonal to their AD or SCZ risks, which potentially unveils resilience and extra-burden factors related to AD or SCZ; 4) benchmark our framework against another method for functional enrichment of genetic risks to evaluate the performance of our framework.

## MATERIALS AND METHODS

### GWAS datasets

The first dataset, Kunkle et al.’s (K) meta-analysis of AD GWAS, focuses on the phenotype LOAD with the age of onset > 65 years, which includes 11,480,632 variants from the study consisting of 21,982 AD cases and 41,944 CN controls [6]. The second GWAS, Schwartzentruber et al.’s (S) meta-analysis of GWAS for AD and AD by proxy phenotypes (GWAX) in the UK BioBank (UKBB), is composed of 898 AD cases, 52,791 AD by proxy cases, and 355,900 controls, including a total of 10,687,077 SNPs [8]. The last dataset is from Wightman (W) et al.’s [7] meta-analysis of AD GWAS. We adopted the summary statistics of their GWAS meta-analysis without the proxy cases from the UKBB and 23andMe, which resulted in 39,918 cases and 358,140 controls, including a total of 12,674,019 SNPs [7]. This design aims to test the robustness and consistency of our parallel subgroup results across various GWAS datasets. SCZ GWAS summary statistics were adopted from Trubetskoy (T) et al.’s meta-analysis, which includes 7,659,767 variants derived from 52,017 SCZ cases and 75,889 controls [19].

### Genotyping data composition in discovery and replication cohorts

We collected AD discovery (disc) and replication (rep) cohorts from dbGaP (https://www.ncbi.nlm.nih.gov/gap/) and synapse (https://www.synapse.org/), respectively. The disc cohort is comprised of the National Institute on Aging/Late Onset Alzheimer’s Disease Study (NIA/LOAD) cohort consents 1 and 2 (ADc12) [phs000168] [39] and the Multi-Site Collaborative Study for Genotype-Phenotype Associations in Alzheimer’s Disease (GenADA) [phs000192] [40]. Raw SNP arrays were downloaded from dbGaP accordingly (accessed on 6/15/2021). The rep cohort includes the Whole Genome Sequencing (WGS) (https://www.synapse.org/#!Synapse:syn5550382) data from the Religious Orders Study and Memory and Aging Project (ROS/MAP) Study [41], the MayoRNAseq (Mayo) study [42], the Mount Sinai Brain Bank (MSBB) study [43], the raw SNP array from Mount Sinai School of Medicine (MSSM) study (https://www.synapse.org/#!Synapse:syn20808201) and imputed SNP array (https://www.synapse.org/#!Synapse:syn3157325) data from ROS/MAP. The raw SNP array from the Alzheimer’s Disease Neuroimaging Initiative (ADNI) database [44] was downloaded from https://adni.loni.usc.edu (accessed on 6/15/2021).

We collected SCZ disc and rep cohorts from the Swedish Case-Control Study of Schizophrenia (SWE) [45] and Molecular Genetics of Schizophrenia (MGS) [phs000167] [46], respectively.

### Imputation of genotyping dataset

To boost the SNPs that are not directly genotyped in the raw genotyping array, we adopted the previous pipeline to impute these raw AD genotyping data [47]. Briefly, we performed the standard variants checking procedure using the tools developed by the McCarthy group [48]. Imputation was conducted for ADc12, GenADA, MSSM, and ADNI datasets using the Michigan Imputation Server (minimac4) [49] with the 1,000 Genome Phase 3v5 European Panel, and SNPs with *r*^2^ < 0.6 were filtered. The imputed genotyping data was downloaded from Synapse, consisting of individuals from the CHOP batch (382) in the ROS/MAP study. From the method description (SynID: syn3157325), the SNPs that passed quality control as described in De Jager et. al [50] were imputed using BEAGLE software, version 3.3.2, with the reference haplotype panels from 87 Centre d Etude du Polymorphisme Humain individuals (CEPH) of Northern European ancestry in the 1000 Genomes Project (1000 Genomes Project Consortium interim phase I haplotypes, 2010-2011 data freeze). For two SCZ cohorts, we adopted the imputation strategy from previous work [51].

### Diagnosis criteria

In this study, we defined AD cases based on the clinical diagnosis labels in each cohort. Specifically, for the AD disc cohort, individuals with dementia who were diagnosed with definitive, probable, or possible Alzheimer’s disease at any stage of their clinical course, according to the criteria introduced in 1984 by the National Institute of Neurological and Communicative Disorders and Stroke and the Alzheimer’s Disease and Related Disorders Association (NINCDS-ADRDA), were categorized as AD cases [52]. Controls were defined as cognitively normal (CN) individuals, excluding those with unspecified dementia, unconfirmed controls, or other neurological diseases.

For the AD rep cohort, we assigned the clinical diagnosis labels to each participant in different sub-cohorts depending on the following criteria. The clinical diagnosis labels were assigned based on cognitive function evaluation and factors available in each cohort. As for the discovery dataset, participants in the ROS/MAP cohort with a consensus diagnosis of AD with no other cause of cognitive function impairment were classified as AD cases, while individuals with no cognitive function impairment were classified as CN controls [53]. Diagnosis in the last follow-up visit was used for clinical diagnosis labeling if a final consensus diagnosis was not available. Individuals in the MSBB cohort were classified based on Clinical Dementia Rating (CDR, range 0 to 5). Participants with CDR ≤ 0.5 were classified as controls, and those with CDR ≥ 3 were classified as AD cases [43]. Participants in the Mayo cohort were classified based on existing clinical diagnosis labels. However, those with a clinical diagnosis of pathological aging or progressive supranuclear palsy diagnosis were removed. For the ADNI and MSSM cohort, AD cases and control diagnoses were based on the last recorded diagnosis status in the metadata.

All SCZ cases in the SWE and MGS cohorts were diagnosed with schizophrenia or schizoaffective disorder (SA) according to the Diagnostic and Statistical Manual of Mental Disorders, DSM-IV or DSM-III-R criteria.

### Genotyping data integration and quality control

We removed non-European ancestry individuals according to their PCA relative to 1000 Genome individuals (**Fig. S1**) [54]. We annotated rsid and European allele frequency for both the disc and rep datasets using ANNOVAR [55] and annotated the rsid column of VCF file using BCFtools [56]. KING2.2 [57] was used to detect relativeness between individuals. We removed individuals with relatives closer than second degree within each dataset. We followed the diagnosis criteria described in the previous section to label the participants and extracted the clinically diagnosed AD and CN individuals for both disc and rep cohorts. We combined the imputed genotyping datasets (ADc12 and GenADA) with VCFtools [58] and formed the disc cohort. Then, we conducted the following variant-based quality control using VCFtools [58], BCFtools [56], and PLINK [59,60], including removing insertion, deletions, and multi-allelic variants; excluding variants without rsid; excluding variants with European allele frequency < 0.01; filtering variants whose call rate were less than 98%; filtering individuals with call rate less than 95%. In addition, we excluded the *APOE* region (chr19, 44.4MB-46.5MB) to create a filtered genotype version for Model 2 of the GRPa-PRS framework, which will be used to impute gene expression without the *APOE* region gene. After the quality control process, the final disc cohort included 2,722 individuals (1,337 cases and 1,385 controls) with 7,232,347 SNPs. Their demographic information and *APOE* genotype of participants are listed in **Table 1**.

**Table 1.** Demographic summary of AD discovery dataset and replication dataset.

|  | Discovery |  |  | Replication |  |  |  |
| --- | --- | --- | --- | --- | --- | --- | --- |
|  | Cases<br>(N=1,337) | Controls<br>(N=1,384) | Total<br>(N=2721) | Cases<br>(N=1,606) | Controls<br>(N=1,248) | Total<br>(N=2,854) | Overall<br>(N=5,575) |
| **Age** |  |  |  |  |  |  |  |
| Mean<br>(SD) | 81.0<br>(7.72) | 75.0 (7.59) | 77.9 (8.23) | 81.4<br>(8.32) | 79.9 (8.75) | 80.8<br>(8.54) | 79.4<br>(8.51) |
| Median<br>[Min,<br>Max] | 81.4 [57.3,<br>103] | 75.0 [57.0,<br>98.0] | 78.0 [57.0,<br>103] | 83.0 [55.0,<br>95.0] | 81.2 [56.0,<br>96.0] | 82.3<br>[55.0,<br>96.0] | 80.0 [55.0,<br>103] |
| **Sex** |  |  |  |  |  |  |  |
| Male | 531<br>(39.7%) | 550<br>(39.7%) | 1081<br>(39.7%) | 586<br>(36.5%) | 578 (46.3%) | 1164<br>(40.8%) | 2245<br>(40.3%) |
| Female | 806<br>(60.3%) | 834<br>(60.3%) | 1640<br>(60.3%) | 1020<br>(63.5%) | 670 (53.7%) | 1690<br>(59.2%) | 3330<br>(59.7%) |
| **APOE**<br>**SNP**<br>**Genotype** |  |  |  |  |  |  |  |
| APOE2/<br>APOE2 | 1 (0.1%) | 6 (0.4%) | 7 (0.3%) | 3 (0.2%) | 7 (0.6%) | 10<br>(0.4%) | 17 (0.3%) |
| APOE2/<br>APOE3 | 42 (3.1%) | 170<br>(12.3%) | 212 (7.8%) | 81 (5.0%) | 164 (13.1%) | 245<br>(8.6%) | 457<br>(8.2%) |
| APOE2/<br>APOE4 | 40 (3.0%) | 28 (2.0%) | 68 (2.5%) | 36 (2.2%) | 17 (1.4%) | 53<br>(1.9%) | 121<br>(2.2%) |
| APOE3/<br>APOE3 | 412<br>(30.8%) | 895<br>(64.7%) | 1307<br>(48.0%) | 723<br>(45.0%) | 808 (64.7%) | 1531<br>(53.6%) | 2838<br>(50.9%) |
| APOE3/<br>APOE4 | 658<br>(49.2%) | 265<br>(19.1%) | 923<br>(33.9%) | 636<br>(39.6%) | 237 (19.0%) | 873<br>(30.6%) | 1796<br>(32.2%) |
| APOE4/<br>APOE4 | 184<br>(13.8%) | 20 (1.4%) | 204 (7.5%) | 127<br>(7.9%) | 15 (1.2%) | 142<br>(5.0%) | 346<br>(6.2%) |

Similarly, we used VCFtools [58] to merge the synapse whole genome sequencing (WGS) [ROS/MAP (1,196), Mayo (346), MSBB(345)], imputed Synapse genotyping dataset [CHOP (382)], imputed MSSM dataset (740) and imputed ADNI dataset (1,065) and formed the rep cohort (SynapseADNI). Among the above participants, only cases and controls were included. We used the same procedure in the disc cohort to conduct the quality control for the genotyping data. The final rep cohort included 2,854 individuals (1,338 cases and 1,154 controls) with 5,970,402 SNPs. Their demographic information and *APOE* genotype are listed in **Table 1**.

For SCZ, the SWE study included 6,628 individuals (2,824 cases and 3,804 controls) with 7,841,039 SNPs, while the MGS study comprised 5,334 individuals (2,681 cases and 2,653 controls) with 7,877,180 SNPs. Their demographic information is listed in **Table 2**.

**Table 2.**
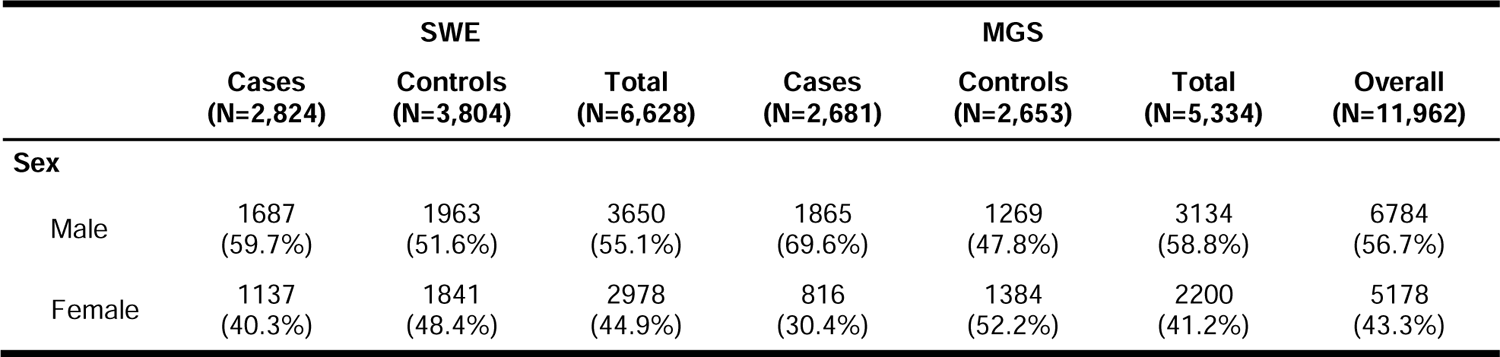
Demographic summary for SCZ SWE dataset and MGS dataset.

### PRS and risk strata comparison

To eliminate inflation caused by sample overlap between the target cohort and GWAS summary statistics in polygenic score analyses, we used the software EraSOR [61] to adjust the effect size from GWAS summary statistics. We used the overlapping variants between the HapMap3 Project [62] and corrected GWAS summary statistics to match the variants in the genotyping data of both cohorts. LDpred2 [63] (-auto model) was applied to calculate PRS for each individual using matched variants. Specifically, we estimated the *h^2^* by LD score regression by ‘bigsnpr’ R package [64], and input *h^2^* and causal variant p (a sequence of 30 logarithmically spaced numbers between 1×10^−4^ and 0.2) into the auto model to calculate PRS. For better classification performance, we repeated the above calculation for different variant p-value thresholds (1, 0.5, 0.2, 0.1, 0.05). For example, when the variant p-value threshold was set at 0.5, only variants with p-values less than 0.5 from the summary statistics were included in the PRS calculation. Their prediction performance was compared using the AUC metrics by using the clinical diagnosis label (Diagnosis criteria section) as the ground truth. The PRS reaching the highest AUC was used to stratify individuals into strata for comparison (**Table 3**). Except for the clinical diagnosis group (Diag) comparison between cases and controls, we repeated the above procedure for three different GWAS summary statistics (Kunkle et al. (K) [6], Schwartzentruber et al. (S) [8], and Wightman et al. (W)) [7] mentioned in the GWAS datasets section and generated three sets with different labels. For SCZ, we used the GWAS by Trubetskoy et al. (T) [19] to calculate the PRS with an optimized p-value threshold of 0.05. We conducted sensitivity analyses using percentiles set at 10%, 15%, and 20% to define strata in both the disc and rep cohorts of AD and SCZ for each comparison.

**Table 3.**
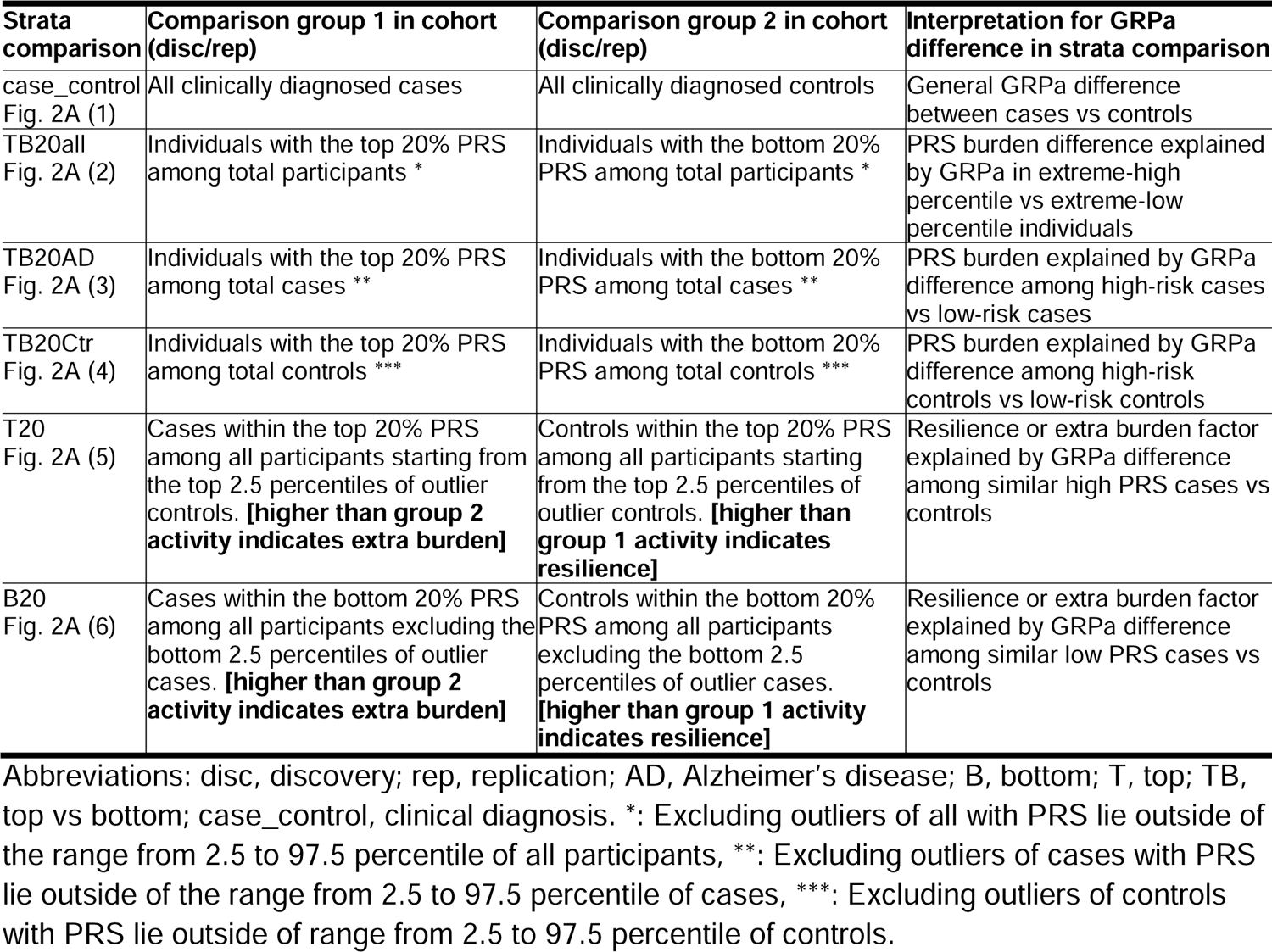
Risk strata and their comparison group interpretation in the discovery and replication data sets.

| Strata comparison | Comparison group 1 in cohort (disc/rep) | Comparison group 2 in cohort (disc/rep) | Interpretation for GRPa difference in strata comparison |
| --- | --- | --- | --- |
| case_control<br>Fig. 2A (1) | All clinically diagnosed cases | All clinically diagnosed controls | General GRPa difference between cases vs controls |
| TB20all<br>Fig. 2A (2) | Individuals with the top 20% PRS among total participants * | Individuals with the bottom 20% PRS among total participants * | PRS burden difference explained by GRPa in extreme-high percentile vs extreme-low percentile individuals |
| TB20AD<br>Fig. 2A (3) | Individuals with the top 20% PRS among total cases ** | Individuals with the bottom 20% PRS among total cases ** | PRS burden explained by GRPa difference among high-risk cases vs low-risk cases |
| TB20Ctr<br>Fig. 2A (4) | Individuals with the top 20% PRS among total controls *** | Individuals with the bottom 20% PRS among total controls *** | PRS burden explained by GRPa difference among high-risk controls vs low-risk controls |
| T20<br>Fig. 2A (5) | Cases within the top 20% PRS among all participants starting from the top 2.5 percentiles of outlier controls. **[higher than group 2 activity indicates extra burden]** | Controls within the top 20% PRS among all participants starting from the top 2.5 percentiles of outlier controls. **[higher than group 1 activity indicates resilience]** | Resilience or extra burden factor explained by GRPa difference among similar high PRS cases vs controls |
| B20<br>Fig. 2A (6) | Cases within the bottom 20% PRS among all participants excluding the bottom 2.5 percentiles of outlier cases. **[higher than group 2 activity indicates extra burden]** | Controls within the bottom 20% PRS among all participants excluding the bottom 2.5 percentiles of outlier cases. **[higher than group 1 activity indicates resilience]** | Resilience or extra burden factor explained by GRPa difference among similar low PRS cases vs controls |

Abbreviations: disc, discovery; rep, replication; AD, Alzheimer's disease; B, bottom; T, top; TB, top vs bottom; case\_control, clinical diagnosis. \*: Excluding outliers of all with PRS lie outside of the range from 2.5 to 97.5 percentile of all participants, \*\*: Excluding outliers of cases with PRS lie outside of the range from 2.5 to 97.5 percentile of cases, \*\*\*: Excluding outliers of controls with PRS lie outside of range from 2.5 to 97.5 percentile of controls.

### Gene set curation

GO curation (Biological Process: BP, Molecular Function: MF, and Cellular Component: CC) is composed of 1,303 non-redundant GO terms from WebGestalt [65] (accessed on 5/7/2021). We obtained the gene set from four different resources, including AD-related pathways and terms and brain cell-type-specific pathway (AD brain function), non-redundant gene ontology (GO), and canonical pathways. We limited the gene number in the gene set ranging from five to 500. Specifically, AD brain function gene sets contain 167 gene sets, curated from two major resources: 70 AD-related function curation [66–68] and 97 brain cell-type-specific functions (https://ctg.cncr.nl/software/genesets (accessed on 6/9/2020) [69–71]). Lastly, we curated 2,082 canonical pathways from three major resources, Kyoto Encyclopedia of Genes and Genomes (KEGG), REACTOME, and BioCarta pathways from the Molecular Signatures Database [72] (MSigDB 7.4 C2 category, accessed on 5/10/2021). Overall, we curated 4202 unique functional gene sets.

### Semantic similarity analysis of GO terms

To better understand the similarities among the significant Gene Ontology (GO) terms identified in different strata, we utilized the R package ‘rrvgo’, which leverages the hierarchical structure of GO terms for BP, MF, and CC separately [73].

### Inferring the GReX for individuals and association study with trait

We used PrediXcan [37] to calculate the GReX via the Multivariate Adaptive Shrinkage in R (MASHR) model [74] for all the individuals using their genotyping data after QC (**Table 1**). The imputed gene expression was calculated for 13 different brain regions from the Genotype-Tissue Expression (GTEx) [75]. These served as the input for two differential GRPa approaches: 1) **GRPa-MAGMA**: TWAS + MAGMA gene set enrichment and 2) **GRPa-GSVA**: gene set variational analysis (GSVA) + logistic likelihood ratio test (LogisticLRT).

### Genetically-regulated pathway-MAGMA gene set enrichment (GRPa-MAGMA)

MultiXcan [38] was first used to integrate TWAS across brain regions and identify the associations between genes and trait. Specifically, MultiXcan performs logistic regression for each gene individually and uses the F-test to assess the significance of the joint fit by comparing the null model (formula (1)) with formula (2). The null model includes only demographic covariates (age, sex, and the top five genotype principal components), while Model 1 includes both demographic covariates and GReX predictors derived from individual genotypes. For each gene, GReX in different tissues was integrated by PCA and treated as predictors in the model. The trait outcome is defined for each stratum using PRS and clinical diagnosis (as shown in **Table 3**).

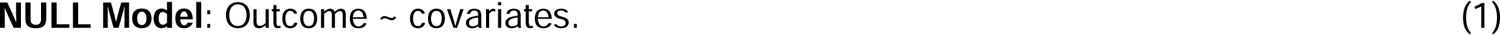

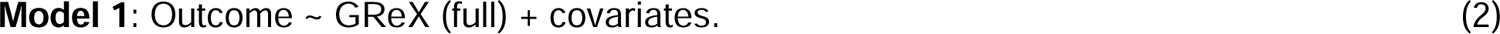

Additionally, due to the dominant effect of *APOE* in AD, we designed Model 2 (no*-APOE* model), which includes both demographic covariates and predictors derived from the genotype excluding *APOE* region as predictors. The GReX of Model 2 was generated by utilizing genotype data that excluded the *APOE* region. The same outcome and covariates were applied in Model 2 as in Model 1. In summary, Model 2 in this study is specifically designed to exclude the genotype or GReX from the APOE region.

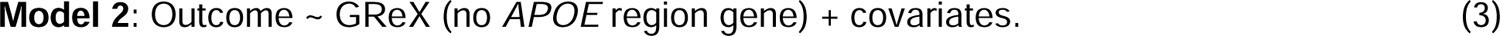

We first employed genotype data of each cohort to construct the gene-gene correlation matrix associated with AD using MAGMA [27]. Based on the results of MultiXcan, we converted the gene p-values to z-scores using the inverse cumulative distribution function (CDF) of the standard normal distribution for a one-tailed test z = Φ^-1^(1 - p) and replaced them with the original z-scores in the MAGMA gene-gene correlation matrix. Then, we adapted the gene set enrichment function in MAGMA with “competitive” and “self-contained” options on three gene set curations, AD brain function, GO, and canonical pathway. We defined a significant GRPa as having a BH-adjusted p-value < 0.05 after applying the Benjamini-Hochberg (BH) test to each gene set curation.

### Genetically-regulated pathway-gene set variation analysis (GRPa-GSVA)

Gene set enrichment was performed for all the imputed AD individuals using the R package GSVA (function gsva - arguments: methodL=L“gsva”, mx.diffL=LTRUE) [76]. GSVA implements a non-parametric unsupervised method of gene set enrichment that allows an assessment of the relative enrichment of a selected pathway across the individual space in a single-sample manner and increases the power to find differential associations [77]. The scores of each GSVA pathway from all individuals followed an approximately normal distribution, which depicted the magnitude difference between the largest positive and negative random walk of each term or pathway. GSVA is a kernel-based method that is less affected by gene-set length or inter-gene correlation [78,79]. We calculated three gene set curations (with minimum five genes and maximum 500 genes) by using the GReX for each individual and each of the 13 brain regions. We proposed two models as GRPa-MAGMA models to detect the conditions with or without *APOE* region GReX. We used logisticLRT to assess the increment in the goodness of fit (deviance of NULL model – deviance of Model 1) and (deviance of NULL model - deviance of Model 2) for each GRPa in each tissue by the Model 1 and the no*-APOE* Model 2, where outcome and covariates were the same as mentioned in the previous model, and GRPa was calculated from GSVA for each tissue and each individual from GReX input.

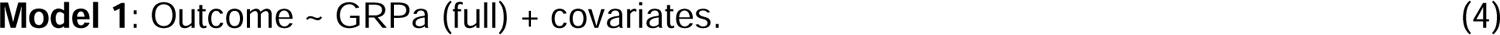

We defined the significant GRPa after Bonferroni correction for each gene set curation.

### PRSet pathway polygenic risk score analysis

To evaluate the performance of our GRPa-PRS, we benchmarked against gene set PRS estimated by one of the latest methods, PRSet [28]. Briefly, PRSet employs the classical approach of clumping and thresholding (C+T) to compute personalized PRSs for specific genomic gene set of interest using individual genotyping information. SNPs within a 35kb upstream and 10kb downstream window of each gene coordinate from the curated gene set were accumulated. To evaluate the association between each pathway and AD outcome, a logistic regression model was applied with the same covariates as GRPa-PRS methods; random SNPs were permuted 10,000 times as the background to assess the empirical competitive *p*-values. We used the BH test to correct for multiple testing in gene sets. The number of pathways with an FDR < 0.05 was used to assess the performance of GRPa-PRS and PRSet across three curated gene sets in two AD cohorts. We conducted logistic regression for genotyping data with and without SNPs in the *APOE* region (Model 1/Model 2) to evaluate PRSet’s performance independent from the *APOE* region, applying the same outcomes and covariates as in previous models.

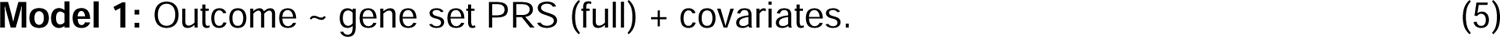

### Comparing the variance explained by GRPa-GSVA and PRSet

To compare the variance explained by the pathway score from GSVA and pathway score from PRSet, we evaluated the goodness of fit in the logistic regression model by Nagelkerke’s R squared based on formula (4) and (5) for each gene set in function curation, using strata case_control as the outcome. In addition, we used both as predictors to compare with formula (5) to explore the improvement that comes from pathway score by our GRPa-GSVA method as shown in formula (6). The same outcome and covariates were applied as the previous models. Furthermore, we divided Nagelkerke’s R squared of the above models by the null model to obtain the R squared ratio to assess the goodness of fit improvement from the generated pathway score.

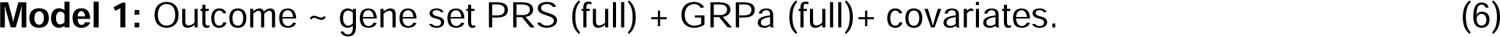

### Orthogonality test to access potential resilience and extra-burden factors

We defined the orthogonal effect as the differential GRPa that does not have a significant correlation with the PRS score. To differentiate GRPas potentially containing resilience and extra-burden factors, we first conducted a Pearson correlation test to determine if the GRPas are significantly correlated with the PRS. Subsequently, we examined the GRPa signal by t-test between all corresponding strata comparisons (**Table 3**). Specifically, the resilience and extra-burden factors are defined by the significantly differential orthogonal GRPas that are identified exclusively in T20 or B20 with no significant difference for case_control or TB20all.

## RESULTS

### Framework of GRPa-PRS and six PRS risk strata comparisons

Individuals carry different genetic liabilities to certain traits and diseases. However, which and how genetic risks play their roles in the pathway and individual level has not been fully revealed. The PRS-based methods have been widely used to characterize such liabilities. We designed a computational framework (**Fig. 1**), genetically-regulated-pathway-polygenic risk score (GRPa-PRS), that can assess the individual-level gene set risk from GReX and further compare the GRPas among biologically meaningful PRS risk strata (**Fig. 2A**). We designed two statistical approaches to identify the differential GRPas and benchmark with variant-based pathway PRS method, PRSet [28].

**Fig. 1.**
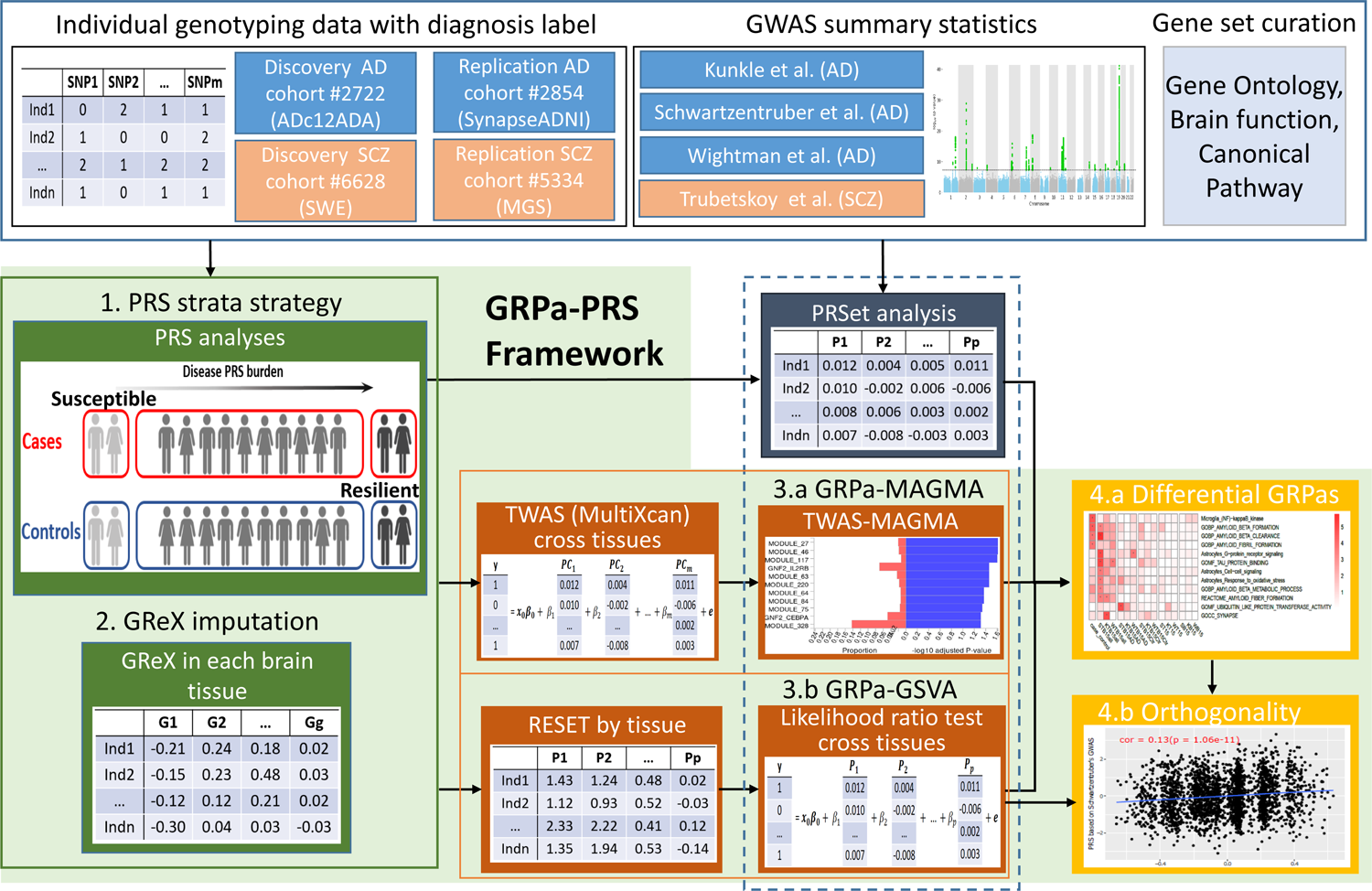
GRPa-PRS workflow and study design. The blue tabulates in the top row indicate the input data from preprocessed individual genotyping data, AD and SCZ GWAS summary statistics, and curated gene sets. The green tabulates and background include the key steps of our GRPa-PRS framework: 1. PRS strata strategy; 2. GReX imputation; 3.a GRPa-MAGMA approach and 3.b GRPa-GSVA; 4.a Differential GRPas summary and 4.b Orthogonality test. The dark blue tabulate includes the method PRSet we used to benchmark the performance of differential GRPa. The dashed line indicates the comparison between three approaches, PRSet, GRPa-MAGMA, and GRPa-GSVA. The orange tabulates are designed to explore the differential GRPa from three different approaches and their orthogonality. The highlighted two strata in Step 1: The resilience stratum (high-risk controls) and the extra-burden stratum (low-risk cases) are defined in **Table 3**.

**Fig. 2.**
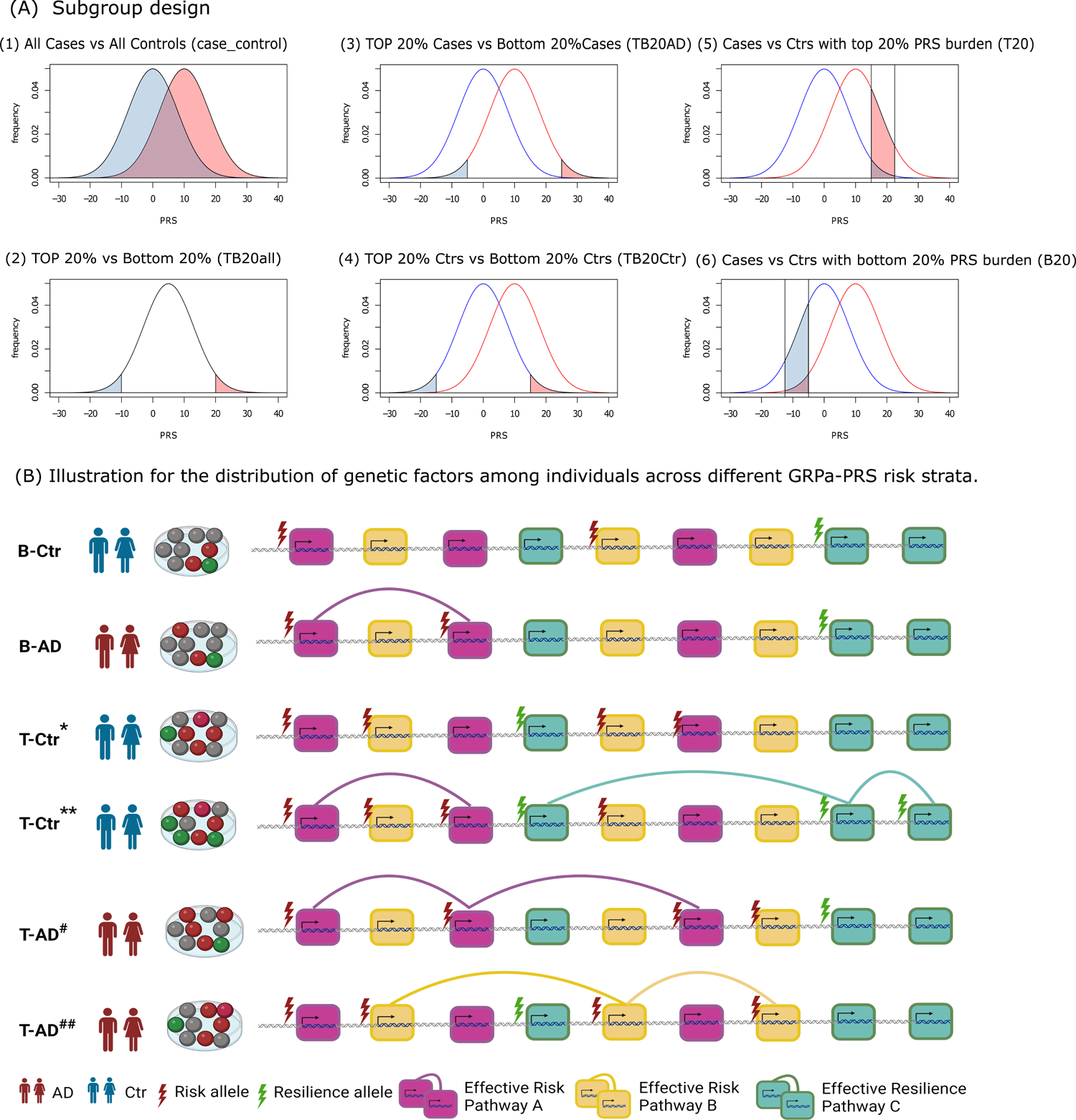
Illustration of six strata comparisons and genetic factor distribution in GRPas. (A) Stratify individuals based on PRS to evaluate underlying differential GRPas as shown in **Table 3**: (1) case_control, (2) TB20all, (3) TB20AD, (4) TB20Ctr, (5) T20, (6) B20. (B) Illustration of the distribution of genetic factors among individuals across different risk strata is shown in **Table 3**. B-Ctr indicates bottom percentile controls carrying no effective risk GRPa. B-AD indicates bottom percentile AD carrying effective risk GRPa. T-Ctr* indicates extreme percentile of controls carrying the risk factors that are sporadically distributed and have no effective risk GRPa; T-Ctr** indicates extreme percentile of controls carrying both effective risk and resilience-related GRPa. T-AD* indicates extreme percentile of cases carrying the risk factors gathered in effective risk GRPa. T-AD** indicates extreme percentile of cases carrying the risk factors gathered in another effective risk GRPa.

GRPa-PRS is designed for polygenic disease in general. In this study, we employed AD and SCZ cohorts to assess the generalizability of the GRPa-PRS framework, with a focus on AD due to its unique nature encompassing two models: one including/excluding the effect of *APOE*. For simplicity, we use only AD to demonstrate how our PRS stratification works. First, we adopted the clinical diagnosis as the case and control labels. After, we estimated the AD PRS risk for each individual using the effect size from the previous three large-scale AD GWAS summary statistics [6–8]. Their PRS model was optimized using LDPred2, applying it separately to three different GWAS summary statistics to ensure a less biased estimation. The six PRS strata comparisons were defined in **Table 3** and illustrated in **Fig. 2A**. Intuitively, for case_control (all cases vs. all controls) stratum and TB20all (top 20% vs. bottom 20%) stratum, we examined GRPa differences among individuals from AD diagnosis groups, focusing on full cases or controls, or extreme percentiles (such 10,15,20) of diagnosis groups (cases or controls). In TB20AD (top 20% cases vs. bottom 20% cases) stratum and TB20Ctr (top 20% controls vs. bottom 20% controls) stratum, we assessed high-risk vs. low-risk burden within AD cases and controls, reflecting the PRS burden within the same diagnosis label. Finally, in T20 stratum comparison (AD cases vs. controls within top 20% PRS) and B20 stratum comparison (AD cases vs. controls within bottom 20% PRS), we explored GRPa differences in individuals with similar PRS burden but different diagnoses, highlighting the counterfactors in resilience and extra-burden-related GRPas.

### PRS model performance evaluation and selection

We calculated the PRS based on three different GWAS summary statistics (S/K/W) adjusted by EraSOR [61] for both disc and rep cohorts. The same PRS calculation process was performed for a list of variant p-value thresholds (1, 0.5, 0.2, 0.1, 0.05). For AD, we found that the prediction performance of different p-value thresholds did not show a large difference. The PRS prediction performance had the highest mean AUC at a variant p-value threshold of 0.2 for both the discovery dataset (AUC S/K/W: 0.69/0.65/0.67) and the replication dataset (AUC S/K/W: 0.68/0.66/0.66). Thus, PRSs calculated at this threshold 0.2 were used to generate the individual PRS, with their distribution shown in **Fig. S2**. The demographic information for each stratum is shown in **Table S1-6.** For SCZ, we used the same process and obtained the highest AUC with a variant p-value threshold at 0.05 for both the disc dataset (AUC T: 0.77) and the rep dataset (AUC T: 0.78). The corresponding demographic information for each stratum is shown in **Table S7-8**.

### *APOE-*region gene sets dominate the differential GRPas in extreme diagnosis groups

As summarized in **Table 3 & Fig. 2A**, we first explored the differential GRPas in PRS burden in the case_control stratum (all cases and controls) and TB20all stratum (top extreme percentile cases and controls vs bottom extreme percentile cases and controls). In the GRPa-MAGMA Model 1 result (**Fig. 3A**), the disc cohort revealed no significant GO terms when comparing cases and controls. Within the three TB20all subgroups, 16 unique significant GO terms were identified. The top three terms were BP amyloid beta metabolic process (FDR: 6.11 × 10^-5^), CC protein lipid complex (FDR: 1.05 × 10^-4^), and BP protein lipid complex subunit organization (FDR: 8.19 × 10^-4^). In the rep cohort (**Fig. 3B**), there are two pathways when comparing cases vs controls and 11 unique significant GO terms within TB20all subgroups, which replicated 7 out of 16 pathways from the disc cohort (**Fig. 3G**). However, for our no*-APOE* Model 2 (**Fig. 3C & 3D**), only one significant pathway (BP amyloid beta metabolic process (FDR: 1.25 × 10^-2^) was identified in the disc cohort and none identified in the rep cohort, suggesting that *APOE*-region genes (**Table S9**) and their related functions dominate the PRS-related GRPas.

**Fig. 3.**
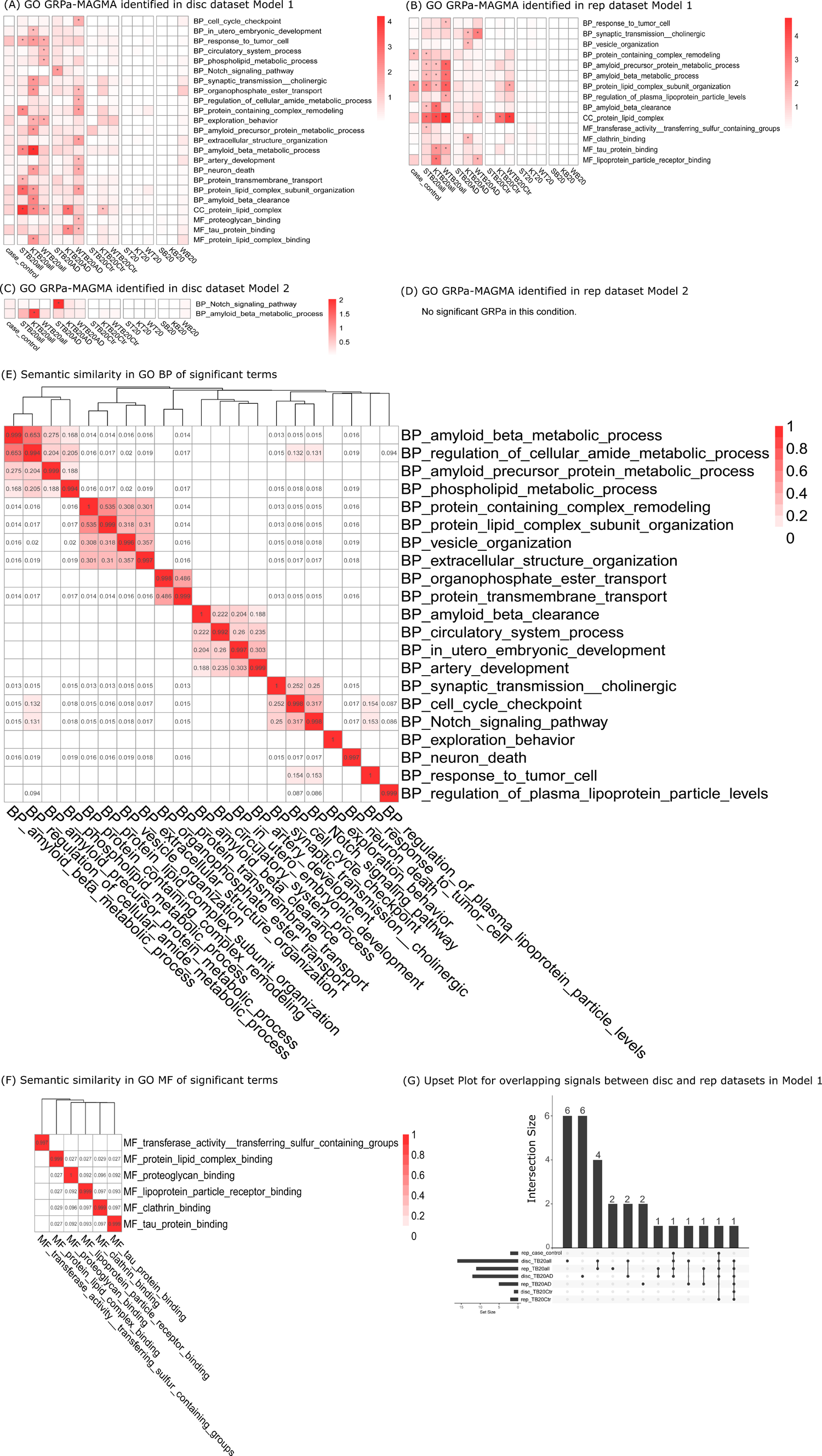
Enrichment of AD GRPa-MAGMA results on GO curation. GRPa was identified by GRPa-MAGMA under Model 1, the model using full genotype to detect all pathways associated with strata comparison, and Model 2, the model using excluding *APOE* region genotype to detect pathways associated with six strata comparisons and independent from *APOE* effect. (A) GO GRPa identified in discovery (disc) dataset Model 1, (B) GO GRPa identified in replication (rep) dataset Model 1, (C) GO GRPa identified in disc dataset Model 2 (no*-APOE* model), and (D) GO GRPa identified in disc dataset Model 2 (no*-APOE* model), no significant result (FDR < 0.05) in this condition. * indicates the significant GRPas FDR < 0.05. Heatmap intensity indicates -log_10_(FDR). The x-axis shows the heatmap list of the subgroup comparison based on different GWAS summary statistics. S represents Schwartzentruber et al; K represents Kunkle et al; W represents Wightman et al.. (E) & (F) show the semantic similarity for significant terms from GRPa-MAGMA in BP and MF, respectively. The UpSet plot for overlapping signals between the strata among the disc cohort and the rep cohort under Model 1 was shown in (G).

In the GRPa-GSVA GO analyses, the disc cohort revealed 14 significant GO pathways when comparing cases and controls (**Fig. 4A**). The top three pathways and terms were BP amyloid beta clearance (p-value: 5.50 × 10^-31^), MF lipoprotein particle receptor binding (p-value: 1.45 × 10^-25^), and BP protein containing complex remodeling (p-value: 2.51 × 10^-17^). Within the three TB20all subgroups, 27 significant GO pathways were identified. The top three terms were BP response to tumor cell (p-value: 3.39 × 10^-41^), MF lipoprotein particle receptor binding (p-value: 2.14 × 10^-24^), and BP amyloid beta clearance (p-value: 5.62 × 10^-20^). The UpSet plot (**Fig. 4G**) demonstrated partial replication of Model 1 results from the disc cohort (**Fig. 4A**) in the rep cohort (**Fig. 4B**) ranging from 41.3%-50.0%. Consistent with the GRPA-MAGMA findings, Model 2 revealed a limited number of pathways (**Fig. 4C & 4D)**. In the discovery cohort, within the TB20all subgroups, amyloid beta-related function was identified even in *APOE-*removed Model 2. Conversely, in the rep cohort, MF clathrin binding (p-value: 8.61 × 10^-6^) and BP vesicle cargo loading (p-value: 3.42 × 10^-5^) emerged only in Model 2, suggesting those terms were independent from the *APOE* effect.

**Fig. 4.**
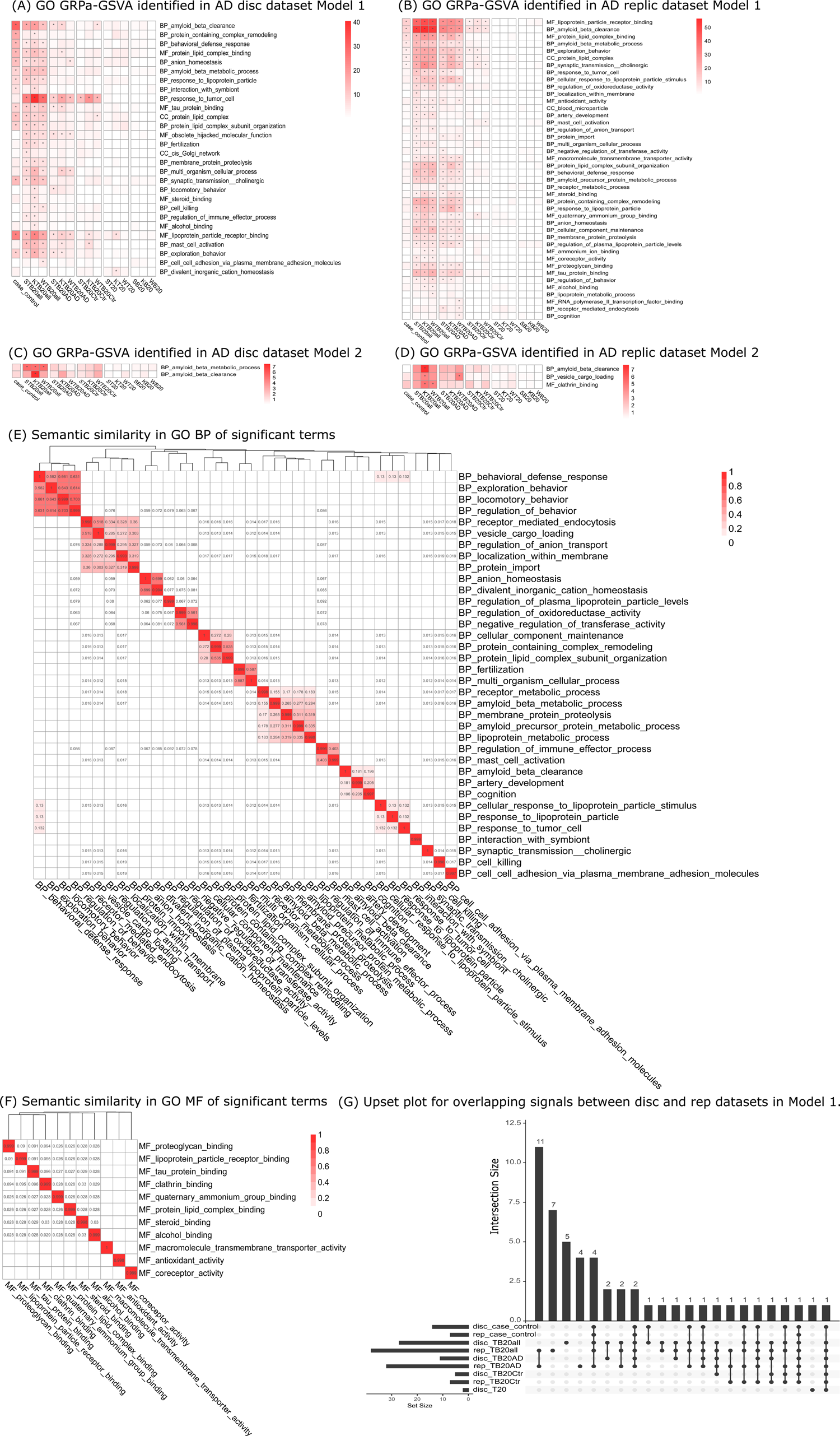
AD GRPa-GSVA results enrich GO pathways. GRPas identified by GRPa-GSVA under Model 1, the model using full genotype to detect all pathways associated with strata comparison, and Model 2, the model using excluding *APOE* region genotype to detect pathways associated with six strata comparisons and independent from *APOE* effect. (A) GO GRPa identified in discovery (disc) dataset Model 1, (B) GO GRPa identified in replication (rep) dataset Model 1, (C) GO GRPa identified in disc dataset Model 2 (no*-APOE*), and (D) GO GRPa identified in disc dataset Model 2 (no*-APOE*). * indicates the significant (p-value < 0.05 / # of gene set) GRPas identified in this condition. Heatmap intensity indicates -log_10_(p-value). The x-axis shows the heatmap list of the subgroup comparison based on different GWAS summary statistics. S represents Schwartzentruber et al; K represents Kunkle et al; W represents Wightman et al.. (E) & (F) show the semantic similarity for significant terms from GRPa-GSVA in BP and MF, respectively. The UpSet plot for overlapping signals between the disc cohort and rep cohort under Model 1 was shown in (G).

We expanded our analysis to include two additional gene set curations (AD brain function terms and canonical pathways, see Methods) by GRPa-MAGMA (**Fig. S3 & S5**), as well as GRPa-GSVA (**Fig. S4 & S6**). The results exhibited a similar pattern to the GO pathways analysis, wherein a limited number of pathways were identified in the no*-APOE* Model 2 compared to Model 1 and TB20all stratum (top extreme percentile cases and controls vs bottom extreme percentile cases and controls) had much more significantly differential GRPas identified than the case_control stratum, highlighting the prominent impact of *APOE* and importance of stratifying individuals to strata based on their PRS.

### TB20AD has more PRS-related GRPas than TB20Ctr does despite their similar PRS burden difference

In the GRPa-MAGMA analysis of GO terms for the TB20AD stratum (high-risk cases and low-risk cases), we observed 12 and five significantly differential GRPas in the disc (**Fig. 3A**) and rep cohorts (**Fig. 3B**). For TB20Ctr stratum (high-risk controls and low-risk controls), there are only two pathways identified in the disc and rep cohorts (**Fig. 3C & 3D**), namely protein lipid complex and protein lipid complex subunit organization. Only protein lipid complex is shared between the disc and rep cohort in GRPa-MAGMA analysis (**Fig. 3G**). Relatively fewer significant GRPas were observed in TB20AD compared to TB20All, yet TB20AD showed more significant GRPas than TB20Ctr. These trends were consistent across results from other gene-set curations. (**Fig. S3 & Fig. S5**). Notably, the TB20AD and TB20Ctr strata shared the same PRS burden difference (**Fig. 2A** panel (3) & panel (4)), which indicates that genetic risk differences in top extreme percentiles of controls are more sporadic and less concentrated in a single pathway (**Fig. 2B**). In our no*-APOE* model, we only identified significant GRPas in TB20AD stratum, including BP Notch signaling pathway (FDR: 9.34 × 10^-3^) (**Fig. 3C**), astrocyte and inhibitory synapse functions in AD brain function gene set curation (**Fig. S3C & S3D**) and REACTOME signaling by notch (FDR: 3.92 × 10^-2^) in canonical pathway gene set curation (**Fig. S5C**).

In the GRPa-GSVA analysis of GO terms for the disc cohort (**Fig. 4A**), we found 11 significantly different GRPas in comparisons within the TB20AD stratum with seven GRPas (19.4%) overlapping with 32 rep cohorts (**Fig. 4B & 4G**). MF RNA polymerase II transcription factor binding and BP cognition were the two significant GRPas that only existed in TBAD20 stratum for GO terms (**Fig. 4B**). For TB20Ctr stratum, we identified the BP myelin maintenance in both STB20Ctr and WTB20Ctr strata in AD brain function gene set curation (**Fig. S4B**) and REACTOME assembly of active LPL and LIPC Lipase complexes (**Fig. S6A**) and KEGG calcium signaling pathway (**Fig. S6B**) in canonical pathway gene set curation. In the no*-APOE* Model 2 for TB20Ctr strata comparison, microglia cell death & apoptosis (**Fig. 4C**) was identified as significant GRPas in AD brain function and KEGG calcium signaling pathway (**Fig. S6B**) in canonical pathway curation, respectively.

### Comparisons in T and B strata highlight potential resilience-related and extra-burden-related GRPas that differ in PRS-matched strata

Lastly, we assessed the potential differential GRPas in PRS-matched strata: T20 stratum (AD cases vs. controls with top PRS) and B20 stratum (AD cases vs. controls with bottom PRS), suggesting the potential resilience and extra-burden factors (counterfactors), respectively. In the GRPa-MAGMA analysis for Model 1 (**Fig. 3A**), only the astrocyte immune system (FDR: 4.84 × 10^-3^) (**Fig. S3A**) and REACTOME in cargo recognition for clathrin-mediated endocytosis (FDR: 3.36 × 10^-2^) (**Fig. S5B**) exist only in T20 stratum. In the no*-APOE* model, the REACTOME pathway cargo recognition for clathrin-mediated endocytosis (FDR: 3.36 × 10^-2^) (**Fig. S5B**) is exclusively identified in the T20 stratum and does not appear in any other PRS strata, suggesting that this pathway may operate independently of *APOE-*related functions.

In the GRPa-GSVA GO analysis for GO the BP divalent inorganic cation homeostasis (**Fig. 4A**) was detected in the disc cohort. KEGG calcium signaling pathway (**Fig. S6B**) is significant in the SB20 stratum. Notably, this term is also highlighted in Model 2 (**Fig. S6D**) and the previous WTB20Ctr stratum, which suggests this differential GRPa is independent from *APOE-*related function.

### Mitochondrial-related function was highlighted in SCZ

To further generalize our framework to another polygenic disease, we applied Model 1 to two SCZ cohorts on GO terms. We identify a few GRPas relatively consistent in GRPa-MAGMA and GRPa-GSVA in each cohort. Specifically, we identified MF structural constituent of muscle in the case_control stratum. BP cytokinesis, BP cell surface receptor signaling pathway involved in heart development, and BP protein localization to cytoskeleton were identified in TBSCZ stratum (**Fig. S7A & S7C**). Mitochondria function-related terms were highlighted in the T stratum (**Fig. S7B**). Lastly, BP translational elongation, MF phosphatidylinositol 3 kinase binding, and BP muscle tissue development were significant in the B stratum (**Fig. S7A & S7C**). Overall, we observed that GRPs were highlighted for their potential roles in muscle and heart development as well as mitochondrial function in T and B strata, indicating their extra-burden or resilience factors.

### Shared and unique significant pathways and terms are identified by three approaches

We assessed the performance of our GRPa-PRS by comparing it to PRSet, a method that computes pathway PRSs based on variants and personalized gene sets for each individual. Within GO, there is a significant difference between the signals identified by the AD disc and rep cohorts. The disc cohort primarily identifies signals in the T20 and B20 strata, whereas the rep cohort signals exist in the TB20all, TB20AD, and TB20Ctr strata (**Fig. S8**). We observed the same discrepancy between these two cohorts within the other two enrichment curations (**Fig. S9 & S10**). Then, we compared the union set of pathways identified using three GWAS summary statistics in the disc and rep datasets across three different methods. The UpSet plot of GO pathways (**Fig. 5A**) shows that nine of the ten largest intersections consist solely of signals from PRSet, without replication from GRPa-MAGMA or GRPa-GSVA, and most are different from established AD-related pathways. Moreover, PRSet identified a disproportionately high number of pathways that more than 240 of 1,303 (18.5%) total GO terms showed significant differences in strata comparisons of two AD cohorts (**Fig. S8**), raising concerns about the extent to which AD pathogenesis can alter GRPas. Compared with our GRPa-MAGMA and GRPa-GSVA, PRSet shows inconsistent replication performance across different cohorts (**Fig. 5A, S11, & S12**). Therefore, we focused on the results obtained from our GRPa-MAGMA and GRPa-GSVA. For example, in the GO gene set (**Fig. 5A**), we found two overlapping signals between these two methods in the case vs. control comparison, 13 (28.9% to 58.8%) overlapping signals in the TB20all stratum, and seven (19.4% to 43.8%) overlapping signals in the TB20AD stratum. In the TB20Ctr stratum, only one signal overlapped, while there were no overlapping signals with other strata. These findings underscore the comparable results between our two methods.

**Fig. 5.**
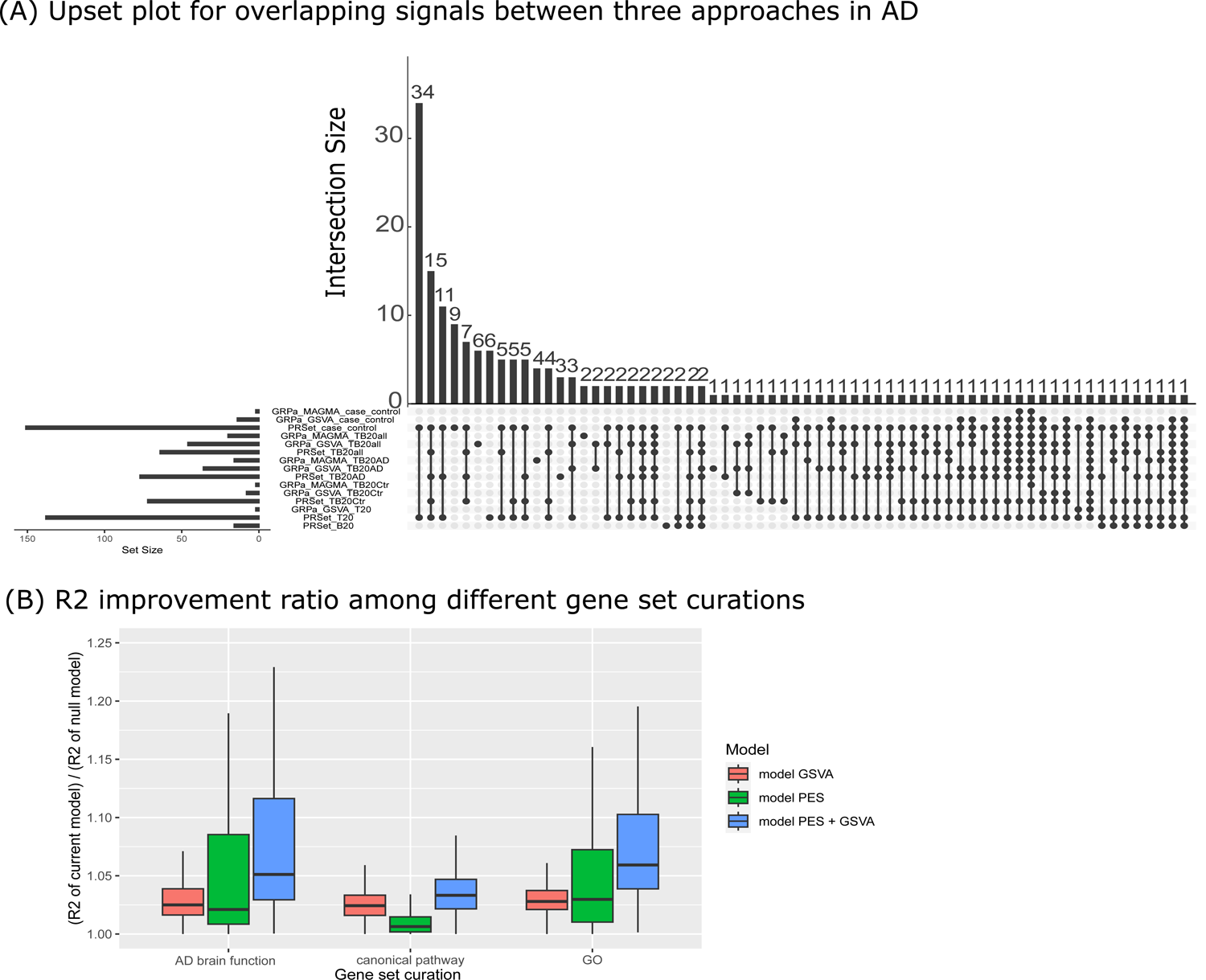
Compare results of three methods GRPa-MAGMA, GRPa-GSVA, PRSet in GReX with *APOE*. The overlapping signals between GRPa-MAGMA, GRPa-GSVA, PRSet are visualized in the UpSet plot (A). (B) shows the R2 improvement ratio using GRPa-GSVA and PRSet from the nested model for formula 4,5,6 among different gene set curations.

### Semantic similarity analyses reveal key functional modules related to significant GO terms

To further summarize our findings with similar functions, we conducted semantic similarity analyses of all significant terms to highlight a few main functional modules (similarity >0.1) shared between GRPa-MAGMA and GRPa-GSVA results for GO terms. Although the terms within each module vary (**Fig. 3E & 4E**), we can still identify major shared terms that cluster into functional modules in BP. These include amyloid beta metabolic processes, amyloid beta clearance, transmembrane and extracellular functions (vesicle, synaptic transmission cholinergic, response to tumor cell, and exploratory behavior). In contrast, MF terms did not cluster into distinct modules; the shared terms are tau protein binding, clathrin binding, proteoglycan binding, and lipoprotein particle receptor binding (**Fig. 3F & 4F**). In the CC category, only protein lipid complex was a shared term between GRPa-MAGMA and GRPa-GSVA results (**Fig. 4G**). Lastly, GRPa-GSVA identified a few unique GO terms, such as anion and cation homeostasis and immune-related functions in GO BP.

For SCZ, BP cell surface receptor signaling pathway involved in heart development from the TBSCZ stratum and muscle tissue development from the B stratum (extra-burden factor) have a shared semantic similarity of 0.25 (**Fig. S7E).** For CC, mitochondrial protein complexes have highly shared semantic functions with other mitochondrial-function-related GO terms from the T stratum with resilience factors (**Fig. S7G)**.

### Gene-level association in significant GRPas from strata comparisons

To further explore the genes that contribute to significant terms, we checked the MultiXcan gene-level p-values and GWAS gene-level p-values. For GRPa-MAGMA, we used the MultiXcan gene-level p-values to reflect the GReX difference in strata comparisons. In AD, the top 20 associated MultiXcan genes (BH-adjusted p < 0.05) (**Fig. S13A & S13B**) were positively correlated with the number of significant GRPas in each strata comparison (**Fig. 3A & 3B**). Ten genes were shared between the top 20 associated MultiXcan genes in AD disc and rep cohorts, including *BIN1*, *PICALM*, and *CD2AP* [80]. In SCZ, no significant MultiXcan genes were identified (**Fig. S13C & S13D**), while well-known SCZ-risk genes [81], such as *ANK3*, *DRD2*, *GRM5*, and *KIF13A* were captured among top genes of either SWE or MGS cohorts.

As GRPa-GSVA does not provide a gene-level association with the trait, we used the GWAS gene p-values derived from MAGMA (Wightman et al. [7] for AD and Trubetskoy et al. [19] for SCZ) to measure the importance of genes from GRPa-GSVA GO terms. For AD, we visualized the top five GWAS significant genes from each significant GRPa across all significant GO terms in AD disc and rep cohorts (**Fig. S14A, S14B & S14C**). As expected, 28 out of 36 GO BP terms included the *APOE* gene (**Fig. S14A**), with the exceptions being immune response, cellular response, and transmembrane activities. For SCZ, we included all the significant GO terms from GRPa-MAGMA and GRPa-GSVA in either SWE or MGS (**Fig. S14D**). Only the mitochondrial function-related terms have shared the top five GWAS significant genes between terms, suggesting that a polygenic effect contributes to SCZ GRPas.

### Comparing the variance explained by GRPa-GSVA vs PRSet

As described in the methods section, we assessed the improvement brought by GRPa-GSVA and PRSet respectively, and jointly by comparing Nagelkerke’s *R* square values of formula (4), (5), and (6) with that of formula (1). The results based on the disc cohort are presented in **Fig. 5B**. Taking the GO gene set as an example, in formula (4), where GRPa generated by GSVA were included as predictors, the ratio of *R* square values ranged from 1.00 to 1.35 (median: 1.03). In formula (5), which includes gene set PRS from PRSet as predictors, the ratio of *R* square values ranged from 1 to 1.61 (median: 1.08). In formula (6), which includes both gene set PRS and GRPa generated by GSVA as predictors, the ratio of *R* square values ranged from 1 to 1.71 (median: 1.11). From **Fig. 5B**, we observed that the improvement in goodness of fit achieved by GRPa-GSVA (formula 4) surpasses that of PRSet (formula 5) in the context of AD brain function and canonical pathways, though it is slightly less effective than PRSet in GO terms. Lastly, it is consistent that adding GRPa-GSVA to PRSet (formula (6)) exhibits significantly better performance compared to PRSet alone (t-test p-value: 1.09 × 10^-7^).

### Definition of orthogonal effect in the resilience and extra-burden GRPas from GRPa-GSVA

In this work, we aimed to identify novel GRPas that are independent from PRS risk and have resilience or extra-burden factors to AD or SCZ, as we defined in the orthogonal effect GRPas session (Methods). Therefore, these resilience and extra-burden GRPas are defined with following features: 1) to be significant in the T20 or B20 strata for resilience or extra-burden factors (**Table 3**); 2) to be orthogonal to the PRS (not significantly correlated with PRS risk). In **Fig. 6**, we illustrated a few pathways and terms as positive and negative examples. For instance, BP divalent inorganic cation homeostasis activity was a notable signal identified in the T20 stratum of the disc cohort. As shown in **Fig. 6A**, there is no significant correlation between its term score and PRS within the brain frontal cortex BA9, suggesting the BP divalent inorganic cation homeostasis activity has an orthogonal effect to PRS liability and is a significant resilience-related GRPa with higher activity in KT20 group 2 than KT20 group 1 (**Fig. 6B**). Similarly, for KEGG calcium signaling pathway, another prominent signal identified in the WTBCtr subgroup and SB subgroup of rep cohort for both Model 1 and Model 2 (**Fig. S6B & S6D**). There is no significant correlation with PRS within the brain spinal cord cervical c−1 (**Fig. 6C**), suggesting the KEGG calcium signaling pathway has an orthogonal effect to PRS liability and is a resilience-related GRPa (**Table 3 & Fig. 2A**) with higher activity in SB20 group 2 than SB20 group 1 (**Fig. 6D**) and higher activity in WTB20Ctrl group 1 than WTB20Ctrl group 2 (**Table S10**). On the contrary, BP amyloid beta clearance, a representative signal identified in both disc and rep cohorts of the case_control, TB20AD, and TB20Ctr strata, but not in the K20 stratum (**Fig. 6F**), demonstrated a significant correlation (*r* = 0.118; p = 6.142 × 10^-10^) with the PRS liability. As depicted in **Fig. 6E**, BP amyloid beta clearance is positively correlated with PRS score and significantly different among the strata comparisons as shown in the heatmap in **Fig. 4A**. For SCZ, BP muscle tissue development demonstrated a non-significant correlation (*r* = −0.021; p = 0.086) with the SCZ PRS liability (**Fig. 6G**), suggesting it is an orthogonal effect. Moreover, BP muscle tissue development has higher activity in stratum B20 group 1 than in B20 group 2 (**Fig. 6H**), indicating extra burdens to SCZ (**Fig. S7C**).

**Fig. 6.**
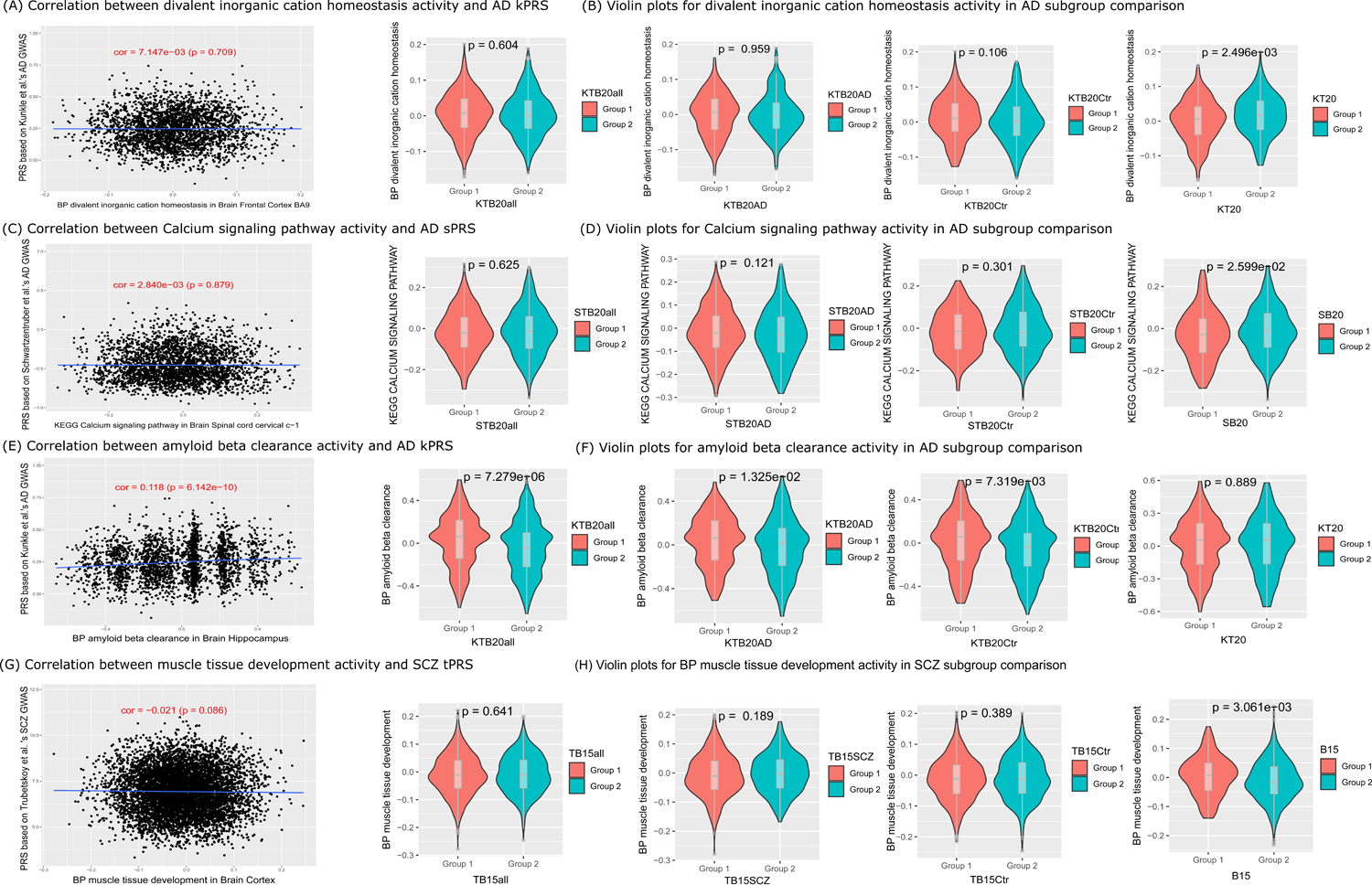
Orthogonal test for key findings. We visualize the correlation between PRS and (A) divalent inorganic cation homeostasis activity, and (C) calcium signaling pathway activity, (E) amyloid beta clearance activity, and (G) muscle tissue development activity, respectively. In (B), (D), (F), and (H), we plot the GSVA score distribution difference within group 1 versus group 2 comparisons among strata comparison, accordingly.

## DISCUSSION

We developed a novel computational framework to explore genetically regulated pathways stratified by PRS (GRPa-PRS) across six predefined risk strata in AD and SCZ, highlighting both well-known and novel gene sets as well as genes responsible for PRS difference among strata. Moreover, we disentangled the GRPas that is orthogonal to, not just correlated to, their AD and SCZ PRS risks, which unveils novel counterfactors playing the roles of resilience and extra burden underlying the risk strata comparison. Notably, we benchmarked our GRPa-PRS method against other variant-based pathway PRS methods and achieved more robust performance, with our framework explaining a greater median proportion of variance.

### Novel risk strata design and robustness

There are a few assumptions for the conjecture’s construction before we can define our risk strata. 1) the diagnosis of each individual mostly reflects their ultimate AD status; 2) the individual disease genetic burden is largely reflected by the PRS; 3) the individuals in each strata comparison might carry different disease genetic risks, but the overall genetic burden is similar (**Fig. 2B**); 4) the individuals are stratified to extreme percentiles (top and bottom 20% here) to reach a maximum PRS disparity as well as a balanced minimum individuals to reach statistical power; 5) considerable proportion of common variant genetic variation can be captured by the variation of GReX genes and their pathway. In other words, the genetic risk difference between strata could be eventually reflected by the GReX difference and functional pathway difference.

With these conjectures, we theorized that different strata, as defined by extremes of PRS load, would provide us with hints as to different resilience and extra-burden genetic factors by identifying those who defy genetic odds by remaining controls despite a high disease genetic load and those who develop disease despite a low genetic load, and also those who conform to genetic odds by remaining controls in concordance to a low disease genetic load and those with who develop disease in concordance to a high genetic load. By systematically contrasting these different strata, our design enabled us to innovatively identify genetic factors associated with resilience and extra burden in our disease models, AD and SCZ. Identifying these genetic factors is the first step toward a framework for enhancing resilience and inhibiting susceptibility, enabling broad preventative efforts.

To ensure the robustness of our findings, we developed two approaches (GRPa-MAGMA and GRPa-GSVA) to systematically characterize the GRPas among risk strata predefined by three GWAS summary statistics and two independent cohorts. Overall, similar trends of GRPa intensity among parallel subgroups were observed (**Fig. 3 & 4**). However, due to the slight difference in each subgroup, significant pathways might not be always shared. Well-known AD-related pathways (including amyloid-beta clearance, tau protein binding, and astrocytes response to oxidative stress) identified by these two approaches are highlighted consistently in case_control, TBall, and TBAD strata in Model 1, suggesting stable GRPas could be observed. Surprisingly, we also identified the amyloid-beta formation and tau protein binding, etc functions significant in Model 2 (no*-APOE*), which suggests other genes such as *BIN1*, *CLU*, *PICALM*, etc could drive the significant difference in GRPas without *APOE*. Besides, GRPa-MAGMA identified astrocyte, neuron, Notch signaling and clathrin functions (**Fig. 3A, 3C, S3C & S3D**), while GRPa-GSVA identified myelin maintenance, clathrin and microglia function (**Fig. 4B, 4D, S4C & S4D**). We observed fewer significant GRPs in the TB20Ctr stratum compared to the TB20AD stratum, despite similar genetic burdens and sample sizes. As illustrated in **Fig. 2B**, this suggests that the additive PRS liability in the top risk controls may result in a more sporadic genetic burden distributed across different GRPas, making them less susceptible than top risk cases.

### Comparison of GRPa-PRS framework with other methods

Despite the lack of ground truth for variant-based gene-set enrichment analysis, we benchmarked the performance of our two GRPa-PRS approaches with PRSet method across three gene set curations in the same strata comparisons of AD disc and rep cohorts. The strong discrepancy between the AD disc and rep cohorts for PRSet among all three gene set curations (**Fig. S8-10**) suggests PRSet is more sensitive to different batches of genotype data. The better performance of our GRPa-PRS approaches may be due to incorporating context-specific gene expression information from disease-related tissues, enhancing signals beyond genotype alone.

### Sample size, effect size and power analysis

In this study, we conducted the PRS calculation with sample sizes of 2,722 and 2,854 for the two AD cohorts, and for SCZ, the sample sizes were 6,628 and 5,334, respectively. The smallest comparison group contains 535 individuals for AD and 534 for SCZ. We expected the effect size of GRPa to have an inverse relationship with the strata size, indicating that when the total sample size is fixed, the effect size (Cohen’s d) increases with higher percentile thresholds. We conducted a sensitivity analysis of different extreme percentiles 10,15, and 20 to explore the relationship between extreme percentile and effect size of terms in supplement figures (**Fig. S16A & S16B**). We could identify a consistent decrease in the absolute median effect size of GO terms across all 13 brain tissues and three parallel PRS-stratification subgroups (K,S,W indicate the GWAS summary statistics used). Consequently, the balance between effect size and sample size (extreme percentile threshold) represents the trade-off between signal and noise. Using the result of **Fig. 4A** as an example, the current GRPas have the largest absolute effect size in BP amyloid beta clearance with an effect size of 0.5 and BP divalent inorganic cation homeostasis with an effect size of 0.25 in TB20AD stratum.

As shown in power analysis at an alpha of 0.05 (**Fig. S16C**), a term with 0.2 effect size in 100 samples will have similar power with 0.15 effect size in 200 samples. All in all, the power analysis and sensitivity analysis further unveil the relationship between GRPa effect size and sample size (extreme percentile). In our current analysis of the extreme 20 percentiles for AD, we anticipate a power of 0.5 to detect effects in strata with a minimum sample size of 200 and an effect size of at least 0.2. For SCZ across the extreme 10, 15, and 20 percentiles (**Fig. S16B**), we expect to achieve the same power, capable of capturing sample sizes ranging from 200 to 500, with minimum effect sizes varying between 0.2 and 0.13.

### Resilience-related and extra-burden-related GRPas interpretation

Through our GRPa-PRS framework, we could identify novel underlying GRPas that contribute to resilience and extra burdens as concluded in **Table 3**. We further defined new resilience-related GRPas with two major criteria: 1) These pathways and terms should be identified as significant in the T or B stratum. Note that these GRPas might also be significant in other strata comparisons, such as TBCtr, as the top risk control stratum might overrepresent resilience-related GRPas. However, these resilience-related GRPas are unlikely to show significant signals in the case-control or TBall strata, where the resilience signal may be diminished within strata comparisons. <u>Therefore, if a GRPa is significant in the T stratum, as well as in case-control and TBall strata, it will not be considered a resilience-related GRPa</u>. 2) The resilience-related GRPas will be further validated using correlation tests to confirm there is no significant association with individual PRS. The criteria could be applied to susceptibility with a different direction of effect.

Overall, we observed two verified resilience-related and one extra-burden-related gene set as well as one potential resilience-related gene set. Firstly, our major findings in GO term explicitly related to resilience were KEGG Calcium signaling pathway (**Fig. S6B**), showing higher activity in SB20 group 2 compared to group 1 (**Fig. 6D**) and in WTB20Ctrl group 1 compared to group 2s (**Table S10**). As advanced age is a major risk factor for AD, age-dependent calcium (Ca2+) dysregulation has been reported in animal studies, showing that aged rodents brains exhibit elevated cytoplasmic Ca2+ ([Ca2+] cyt) and increased overall Ca2+ levels [82,83], Nevertheless, presenilin mutations, accounting for over 90% of familial AD cases, disrupt cytoplasmic and mitochondrial Ca2+ homeostasis, leading to neurodegeneration [84]. Increasing evidence [85] indicates that Ca2+ dysregulation due to presenilin mutations occurs before the formation of Aβ plaques and neurofibrillary tangles in the AD brain. This suggests that disruptions in cytoplasmic Ca2+ may be the primary origin of AD, which aligns with our finding that the genetically-regulated calcium signaling pathway is a key GRPa related to AD outcomes. More importantly, since Ca2+ disruption is central to AD pathology, targeting Ca2+ channels or interfering in proteins with chemical agents or small molecules could offer new avenues for AD prevention and treatment, distinct from approaches based on the amyloid cascade hypothesis [86].

Another term related to resilience, divalent inorganic cation homeostasis activity, (**Fig. 4C, 6C & 6D**) is composed of the activity of several homeostasis of essential biometals (e.g., iron [Fe2+], copper [Cu2+], calcium, manganese [Mn2+], zinc [Zn2+], and magnesium [Mg2+]). The cell surface transporter, such as divalent metal transporter 1 (DMT1), could transport Fe2+, Cu2+, Mn2+ in the cytosol of neurons, astrocytes, and microglia [87]. Two isoforms of DMT1, DMT1-IRE, and DMT1+IRE, colocalize with Aβ plaques in the AD brain, with both isoforms significantly elevated in the frontal cortex and hippocampus of an APP/presenilin transgenic mouse model, suggesting that deregulation of inorganic cation metabolism proteins DMT1 plays a critical role in AD neuropathogenesis [88]. Additionally, the Zn2+ transporter SLC39A13 (ZIP13) and Ca2+ transporter SLC24A4 (NCKX4) are among the top five GWAS risk genes from the divalent inorganic cation homeostasis activity term (**Fig. S14**).

From the GRPa-GSVA of AD brain function gene set, we also identified one potential resilience-related gene set microglia cell death & apoptosis in disc AD TB20Ctr Model 2 (**Fig. S4C**). We identified the orthogonal effect (**Fig. S15A**) and higher activity in group 1 [extreme percentile controls] compared to group 2 [bottom percentile controls] (**Fig. S15B**). Microglia plays a double-edged sword role in AD. Initially, they facilitate the clearance of amyloid-beta (Aβ) and contribute to maintaining brain homeostasis. However, the accumulation of Aβ triggers the release of proinflammatory cytokines, which in turn impairs the microglia’s capacity to effectively clear Aβ. Additionally, activation of the NLRP3 inflammasome releases apoptosis-associated speck-like protein (ASC), which binds to Aβ, causing its aggregation and further spreading amyloid pathology [89]. Therefore, reducing the number of over-activated microglia through apoptosis can reduce the levels of inflammatory mediators, consequently mitigating neuroinflammation and its deleterious effects on neuronal integrity. On the contrary, we also identified a significant GO BP term myelin maintenance in rep AD TB20Ctr Model 1 (**Fig. S4B**). However, we failed to verify the orthogonal effect (**Fig. S15C**), therefore, we can’t determine it as a potential resilience-related GRPa.

Despite three resilience-related pathways and terms in AD, we only identified one significant extra-burden BP GO term muscle tissue development activity in SCZ (**Fig. S7C, 6G & 6H**) with higher activity in B20 group 1 [bottom percentile cases] than in B20 group 2 [bottom percentile controls] (**Fig. 6H**). *FLOT1*, *ERBB4*, *SGCD*, *DDX39B*, and *ALPK3* are the top five most significant genes in muscle tissue development activity (**Fig. S14D**) from SCZ GWAS summary statistics [19]. Specifically, *FLOT1* (Flotillin 1) is involved in lipid raft-mediated signal transduction, which is important for muscle cell differentiation and regeneration [90]. *FLOT1* is involved in neurodevelopmental processes. Alterations in *FLOT1* expression have been linked to neuropsychiatric disorders, including schizophrenia [91]. *ERBB4* (Erb-B2 Receptor Tyrosine Kinase 4) plays a role in both neuromuscular junctions [92] and neural development, impacting synaptic plasticity and neurotransmission [93]. *SGCD* (Sarcoglycan Delta) is part of the sarcoglycan complex crucial for muscle cell membrane integrity. Mutations in SGCD are linked to muscular dystrophies [94]. SGCD was also found in a run of homozygosity locus linked to SCZ [95]. Overall, these genes illustrate how disturbances in developmental pathways can contribute to both muscular and psychiatric abnormalities, highlighting a potential intersection of muscular and neural function in the etiology of SCZ.

### Limitations

First, due to a lack of individual information in the GWAS summary statistics, the overlapping individuals among our two cohorts and GWAS summary statistics could not be detected. This might have led to bias in the PRS calculation. Given the large samples (at most less than 1% of samples are overlapped) in GWAS summary statistics, we believe the impact of this possible bias to be mitigated after adjusting sample overlap using EraSOR [61] method. Indeed, we obtained an AUC of ∼0.67 in AD and AUC of ∼0.77 in SCZ) after the overlap sample correction, which aligns with the AUC performance of ∼ 0.8 in AD [21,96] and ∼0.72 in SCZ [97] from previous studies. Second, by borrowing the expression information from diseases/traits-related tissues, our GRPa-PRS framework has more power to detect GRPa independent from *APOE* at the cost of higher computation burden and time than SNP-based methods like PRSet. Third, although we identified relatively consistent signals in Model 1 (TBall, TBAD, and TBCtr) among three GWAS summary statistics and two independent cohorts using our two approaches in the GRPa-PRS framework, we failed to identify relatively stable signals in Model 2 and other strata (T and B) in either Model 1 or Model 2. One potential explanation for this is the small sample size and small effect size of resilience-related GRPas compared to *APOE-*related GRPas, as expected. Fourth, our newly developed method does not consider the effect of rare variants; this fact might have had a larger impact in the comparisons involving the extra-burden group where rare variants possibly play a larger role due to their higher susceptibility. Fifth, we chose a soft threshold of 20% and met a minimum sample size of 500 in each comparison extreme stratum and simultaneously maximize the difference (**Table S1-S8**). Notably, the results of soft thresholds of 10% and 15% are comparable to 20%, despite the decreasing of the effect size (**Fig. S16A & S16B**). We recognized that a larger cohort would increase our power to detect significant GRPas (**Fig. S16C**). Therefore, applying our model to other polygenic diseases with larger cohorts would benefit from a more stringent percentile threshold, maximizing the effect size of genetic differences. Sixth, we also observe that there is a tendency for more APOE4 alleles in the high PRS group than in the low PRS group. We designed to conduct the differential GRPas comparison in strata without using APOE4 allele dosage as the covariate given the dosage difference listed in **Table S1-S8**. We also stratified the case-control comparison using *APOE* genotype (22,23,33,34,44) of individuals and conducted the GRPa-MAGMA for three curated gene sets. Only one new significant GRPa was identified (**Fig. S17**), suggesting that there are limited significant GRPas between *APOE* genotype stratified case and control comparison, and the significant differential GRPas between case-control comparison are mainly driven by *APOE* genotype dosage differences in case and control groups. Seventh, the GRPa-PRS framework is designed to identify the GRPas among different PRS strata. Therefore, adjusting the PRS value in the regression model will significantly mitigate the genetic signals as shown in **Fig. S18**. The current orthogonal test only allows us to validate the results from GRPa-GSVA. Using GRPa-MAGMA, astrocytes immune system was identified in the ST20 subgroup for AD brain function and might be a strong candidate for an orthogonal effect as it could be identified by both GRPa-MAGMA and GRPa-MAGMA with PRS correction (**Fig. S3A & S18B**). Therefore, using the PRS value as a covariate might be an alternative way to test the orthogonal signals for the T and B strata, which will be explored in future work. Finally, the minimum age of the included controls was 57 years, thus we cannot guarantee that some individuals belonging to the top risk control stratum will not eventually develop AD as they age. However, we believe that this makes our model conservative when searching for resilience factors and that it leads to false negatives but increases the odds that the found signals are indeed true positives. In addition, age of onset as a limitation applies mostly to our exemplar AD and should play a considerably lesser role in other polygenic disorders with development earlier in life.

### Future directions

We used AD and SCZ as prototypes to demonstrate our GRPa-PRS framework, with AD serving as a special case and SCZ representing a more general polygenic disorder. Another exciting possibility is to incorporate into GRPa-PRS the capacity to identify different strata with different resilience and extra-burden factors; this could enable personalized prevention by reinforcing the different resilience factors and weakening the different extra-burden factors of each individual according to their specific PRS strata instead of reinforcing or mitigating them universally in all individuals. Thus, the identification of cluster pathways using GRPa-PRS could guide precise prevention. Finally, the incorporation of the disease strata according to PRS developed in GRPa-PRS to single-cell data could lead to new biological insights not possible using PRS alone.

## Conclusions

We developed a framework, **GRPa-PRS**, including two approaches (GRPa-MAGMA and GRPa-GSVA) to systematically explore the differentially genetically-regulated pathways among individuals stratified by their AD risks, ultimately revealing resilience and extra-burden biological factors. The GReX-level comparison among the strata revealed pathways associated with disease risks, highlighting both well-known and novel pathways and genes. We highlighted one metabolic pathway calcium signaling pathway playing a resilient role independent from PRS risk. This GRPa-based PRS framework can be extended to other polygenic complex disorders and uncover the underlying counterfactors in high-risk controls that allows them to evade susceptibility as indicated by the PRS.

## DATA AVAILABILITY

All the data generated or analyzed in this study is available from the authors upon reasonable request. The overall framework can be downloaded from https://github.com/davidroad/GRPa-PRS.

https://www.ncbi.nlm.nih.gov/projects/gap/cgi-bin/dataset.cgi?study_id=phs000168.v2.p2&pht=710
https://www.ncbi.nlm.nih.gov/projects/gap/cgi-bin/analysis.cgi?study_id=phs000219.v1.p1&pha=2879#:~:text=GenADA%20is%20a%20multi-site%20collaborative%20study%2C%20involving%20GlaxoSmithKline,variations%20in%20candidate%20genes%20with%20Alzheimer%E2%80%99s%20disease%20phenotypes
https://www.synapse.org/#!Synapse:syn5550382
https://www.synapse.org/#!Synapse:syn3157325
https://adni.loni.usc.edu

## ACKNOWLEDGEMENTS

We would like to thank all the members of Bioinformatics and Systems Medicine Laboratory (BSML) for constructive discussion.

## Abbreviations

AD: Alzheimer’s disease;

AUC: area under the receiver operating curve;

BP: Biological Process;

CC: Cellular Component;

CI: confidence interval;

CN: cognitively normal;

CDR: clinical dementia rating;

disc: discovery;

FDR: false discovery rate;

eQTL: expression quantitative trait loci;

GO: gene ontology;

GRPa: genetically-regulated pathway;

GRPa-MAGMA: genetically-regulated pathway-gene-set enrichment analysis;

GRPa-GSVA: genetically-regulated pathway-gene-set variational analysis;

GReX: genetically-regulated expression;

GSEA: gene-set enrichment analysis;

GSVA: gene-set variational analysis;

GWAS: genome-wide association study;

GWAX: genome-wide association study based on proxy phenotype;

*h^2^*: heritability;

KEGG: Kyoto Encyclopedia of Genes and Genomes;

LOAD: late-onset Alzheimer’s Disease;

LogisticLRT: logistic likelihood ratio test;

MF: Molecular Function;

OR: odds ratio;

PRS: polygenic risk scores;

rep: replication;

TWAS: transcriptome-wide association study;

WGS: Whole Genome Sequencing;

Risk strata comparison: T: top;

B: bottom;

TB: top vs bottom;

Ctr: control;

TBall: top vs bottom for all individuals;

TBAD: top vs bottom for only AD patients;

TBCtr: top vs bottom for only controls;

Parallel subgroups: K: stratify by PRS based on Kunkle et al.’s AD

GWAS; S: stratify by PRS based on Schwartzentruber et al.’s AD GWAS;

W: stratify by PRS based on Wightman et al.’s AD GWAS;

T: stratify by PRS based on Trubetskoy et al.’s SCZ GWAS;

## Data source

ADc12: National Institute of Aging/Late Onset Alzheimer’s Disease Study (NIA/LOAD) cohort consents 1 and 2; GenADA: Multi-Site Collaborative Study for Genotype-Phenotype Associations in Alzheimer’s Disease; ROS/MAP: Religious Orders Study and Memory and Aging Project; Mayo: MayoRNAseq; MSBB: Mount Sinai Brain Bank study; MSSM: Mount Sinai School of Medicine study

