## Supplemental Figure 6 for "GRPa-PRS: A risk stratification method to identify genetically-regulated pathways in polygenic diseases"

(A) Canonical Pathway GRPa-GSVA identified in AD disc dataset Model 1

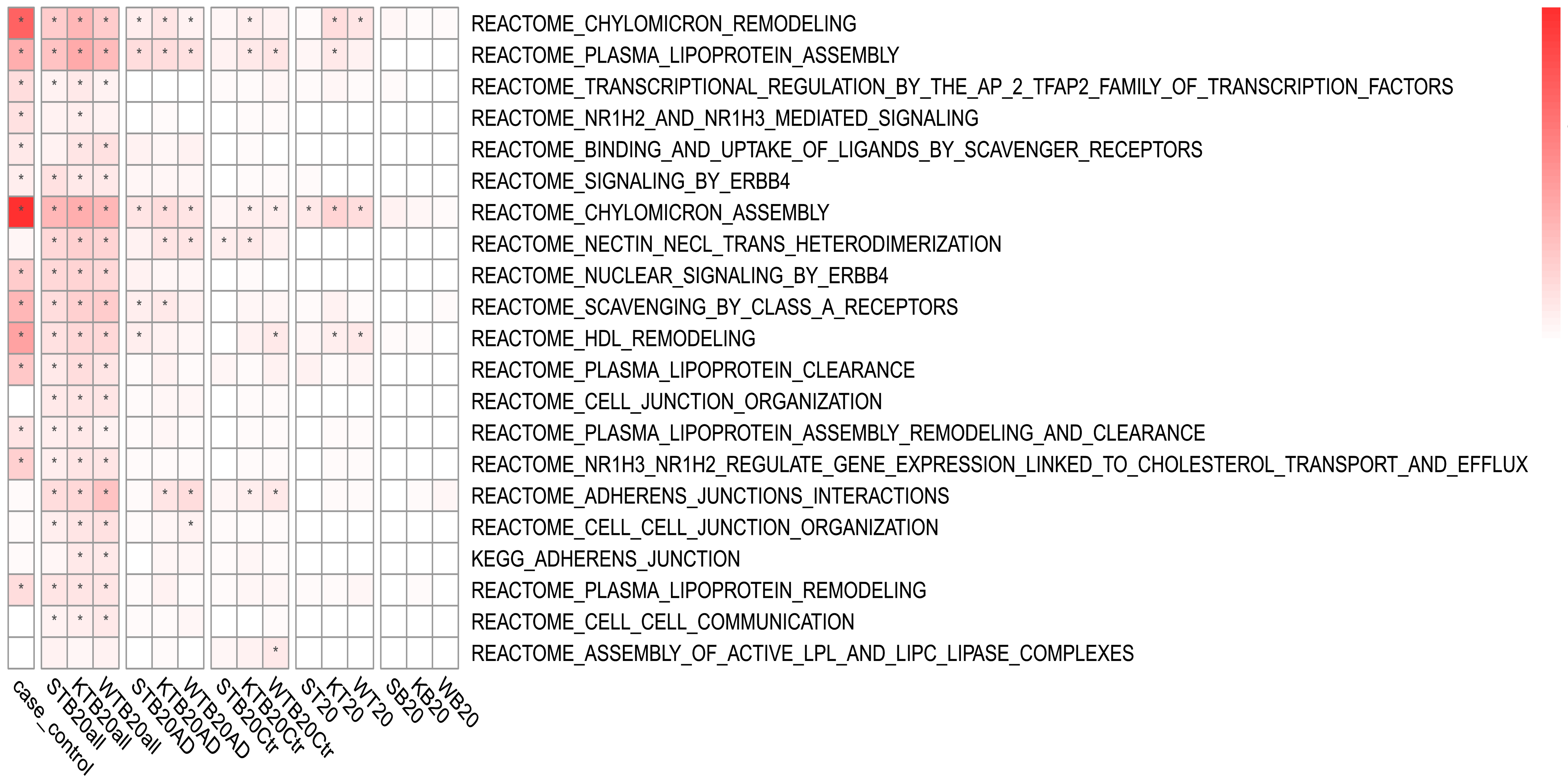


(B) Canonical Pathway GRPa-GSVA identified in AD rep dataset Model 1

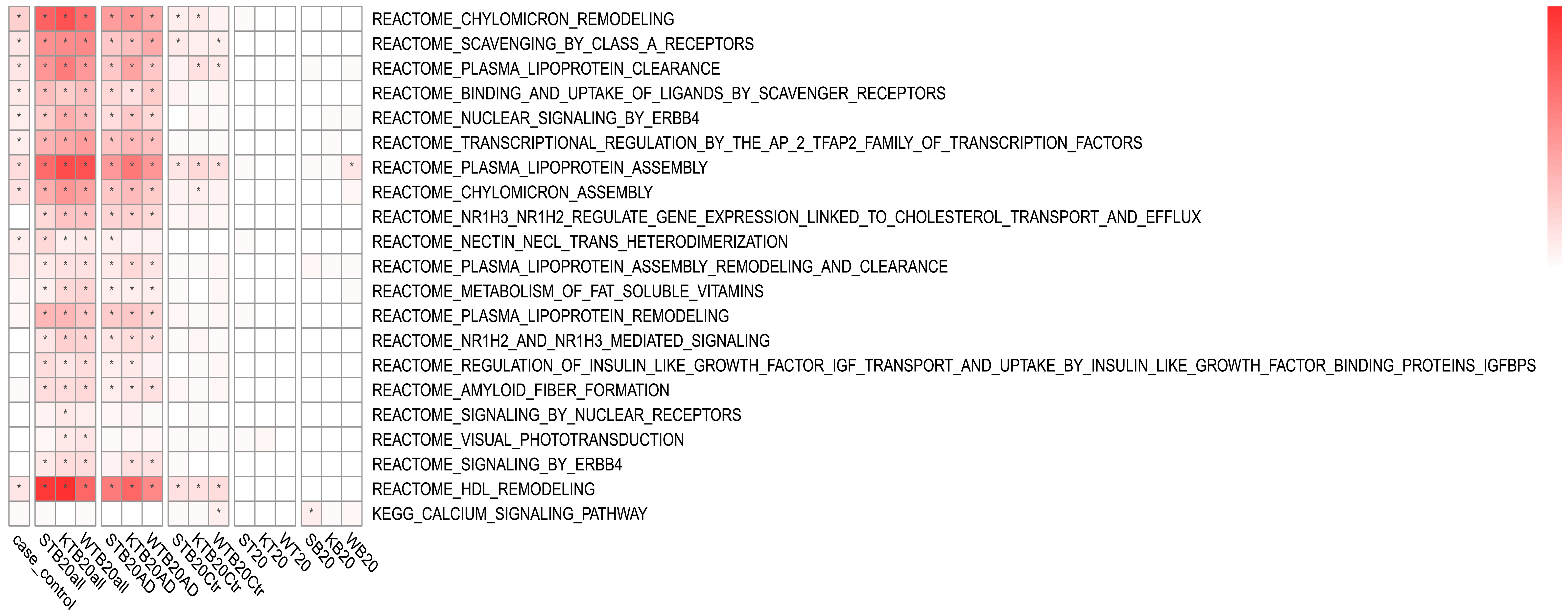


(C) Canonical Pathway GRPa-GSVA identified in AD disc dataset Model 2

No significant GRPa in this condition.

(D) Canonical Pathway GRPa-GSVA identified in AD rep dataset Model 2

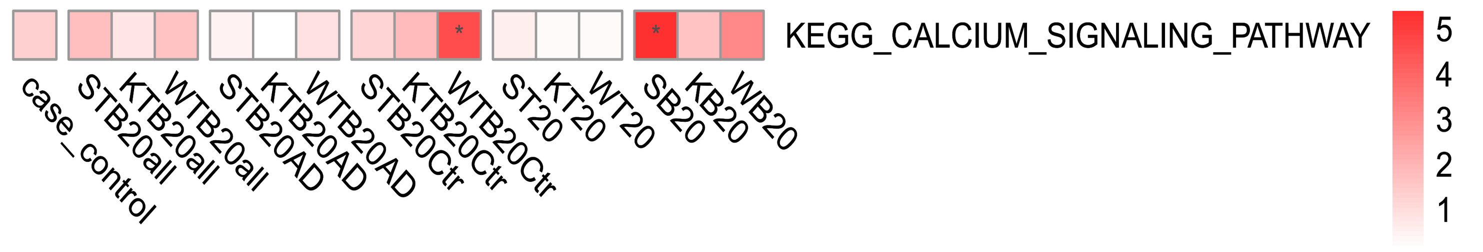


(E) Upset Plot for overlapping signals between dis and rep datasets in Model 1

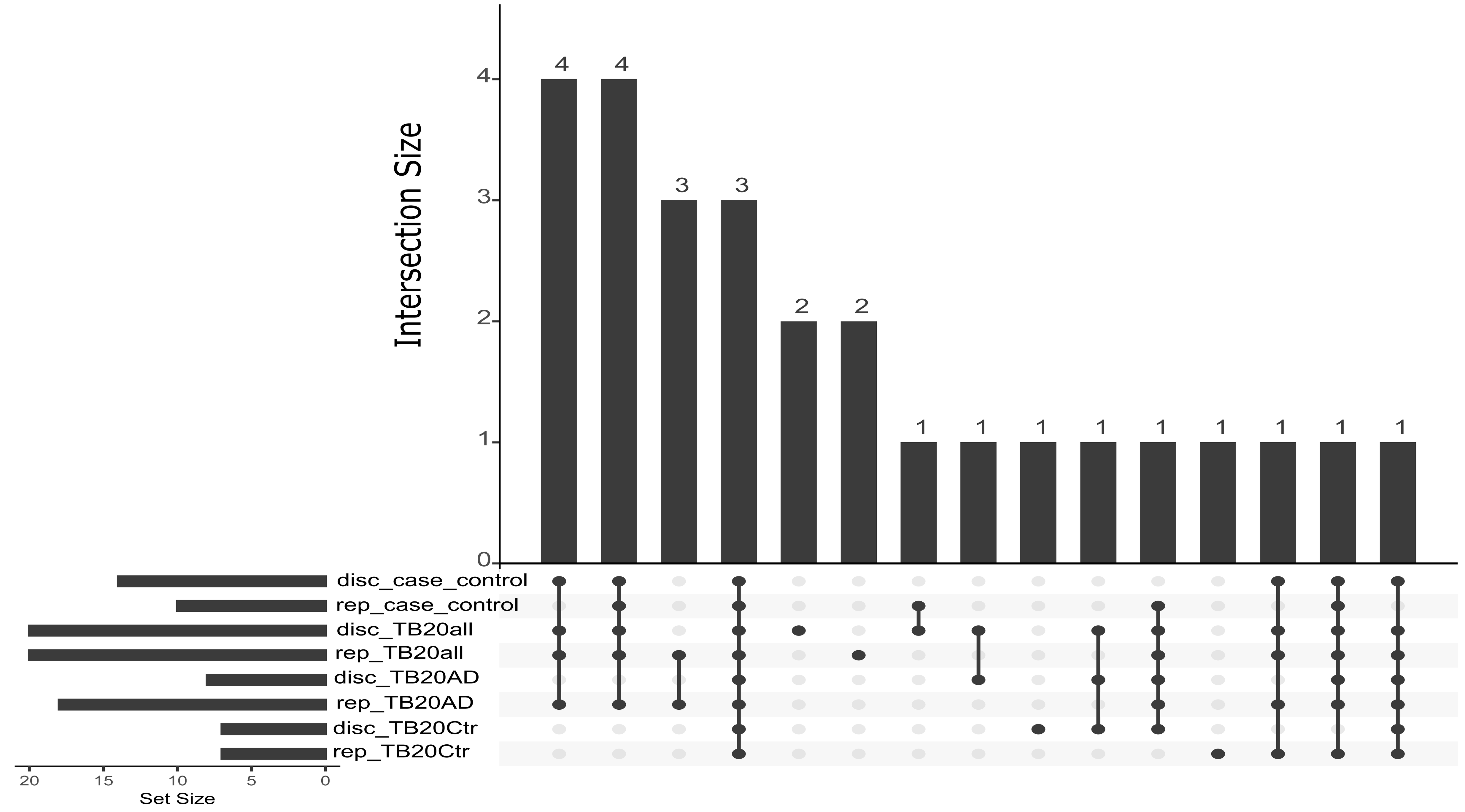
