## Supplemental Figure 8 for "GRPa-PRS: A risk stratification method to identify genetically-regulated pathways in polygenic diseases"

(A) GO PRSet identified in AD disc dataset Model 1

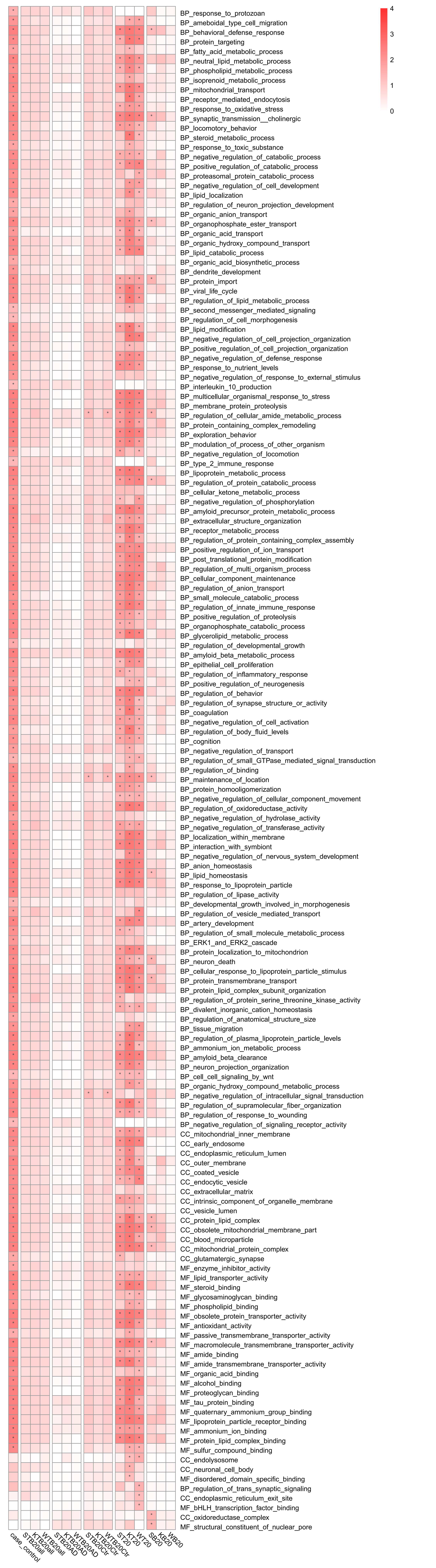

(B) GO PRSet identified in AD rep dataset Model 1

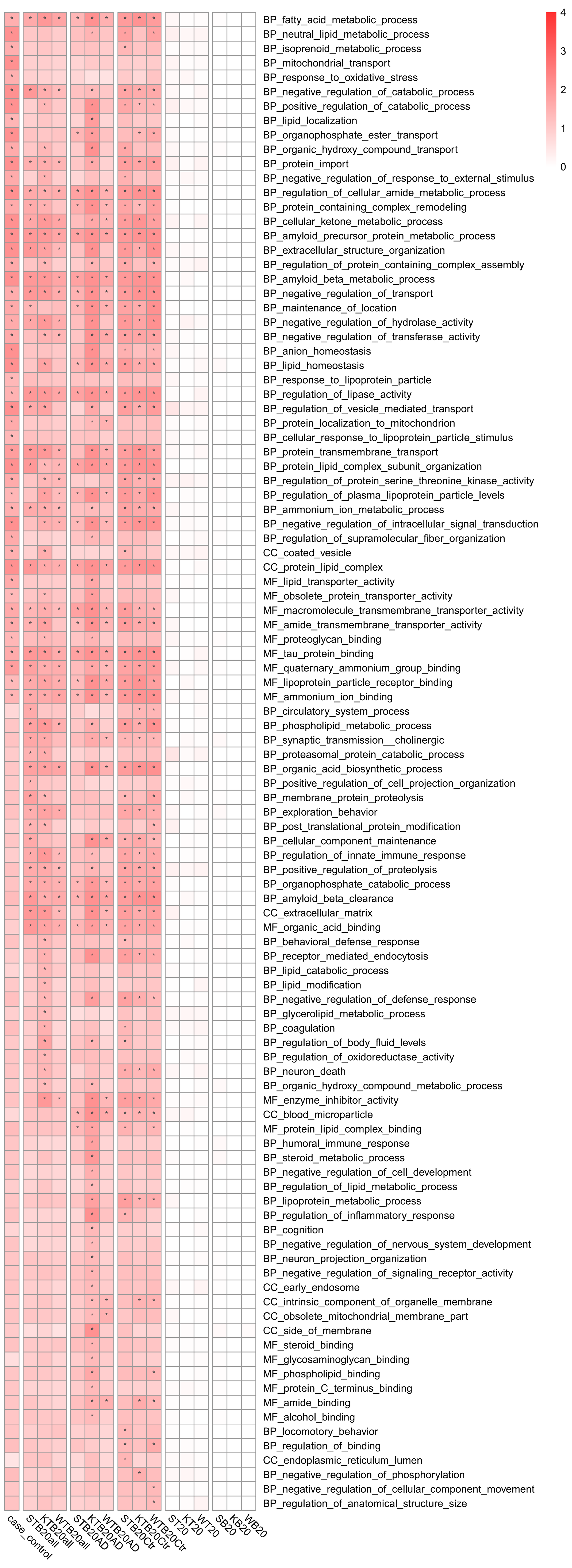

(C) GO PRSet identified in AD disc dataset Model 2

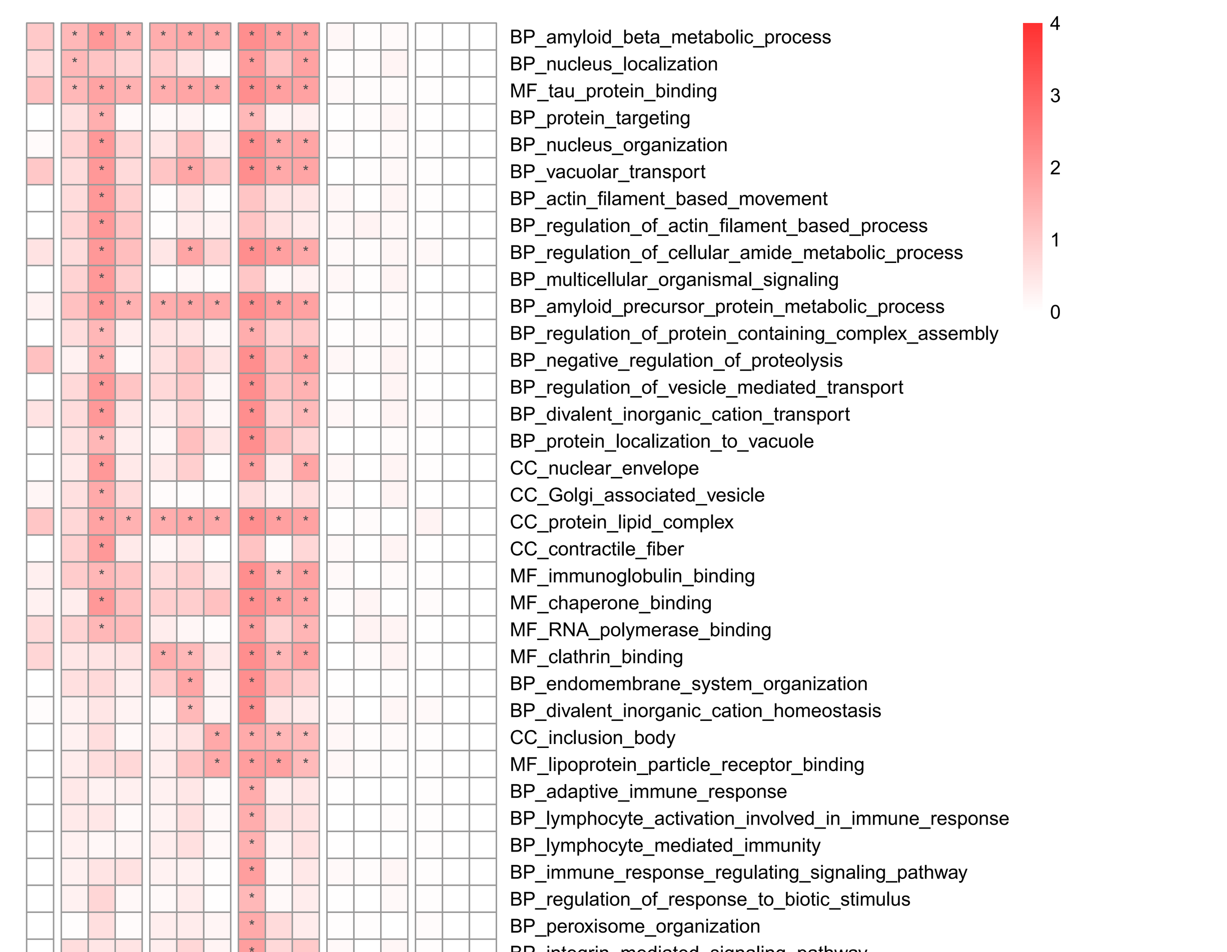

(D) GO PRSet identified in AD rep dataset Model 2

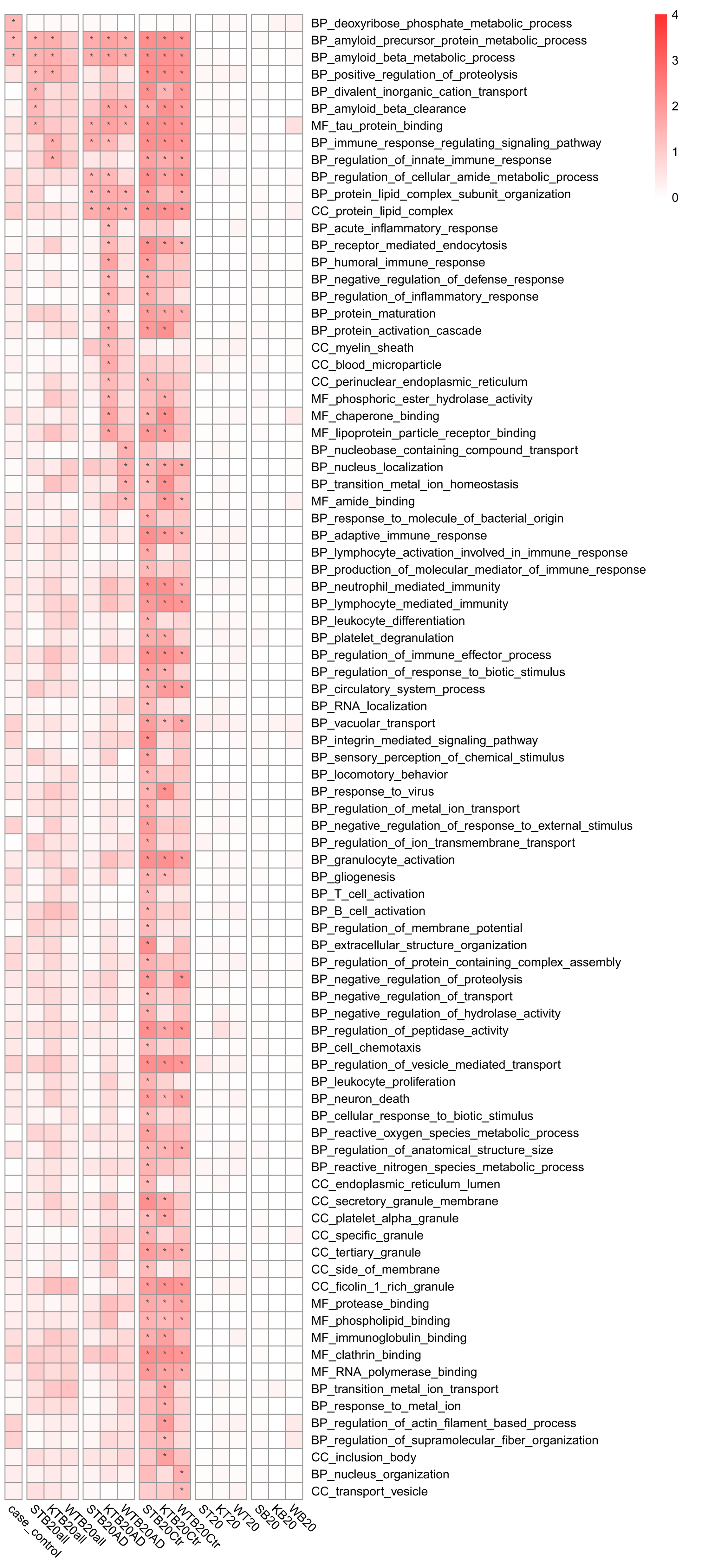

(E) Upset plot for overlapping signals between disc and rep datasets in Model 1.

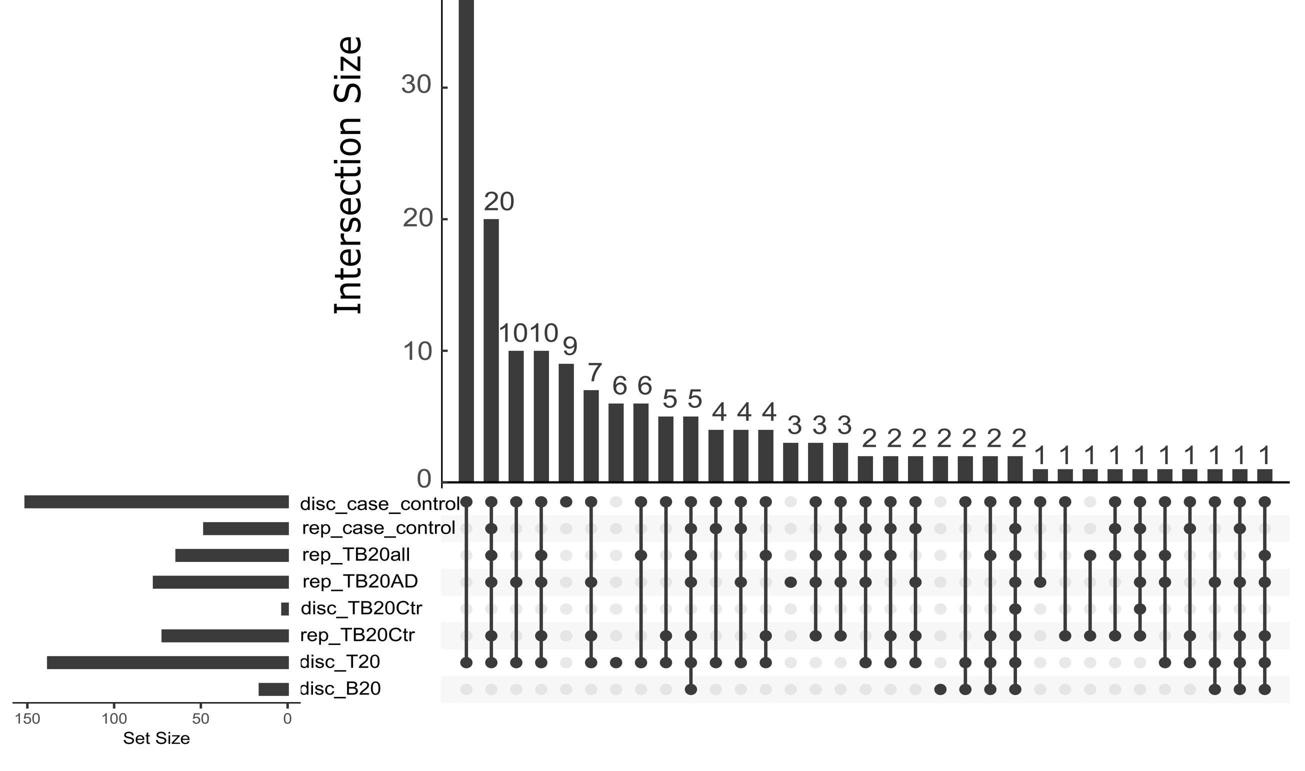
