## Supplemental Figure 10 for "GRPa-PRS: A risk stratification method to identify genetically-regulated pathways in polygenic diseases"

(A) Canonical Pathway PRSet identified in AD disc dataset Model 1

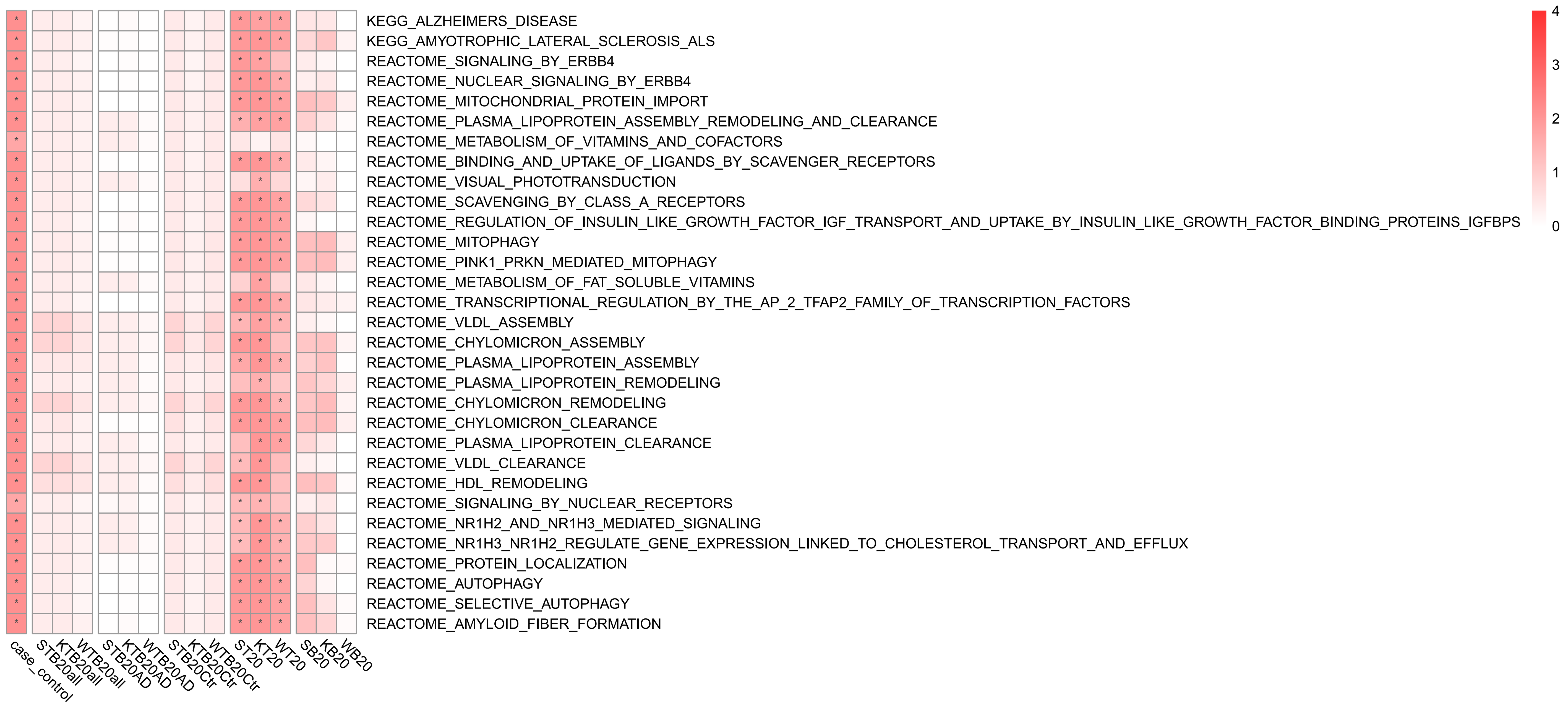


(B) Canonical Pathway PRSet identified in AD rep dataset Model 1

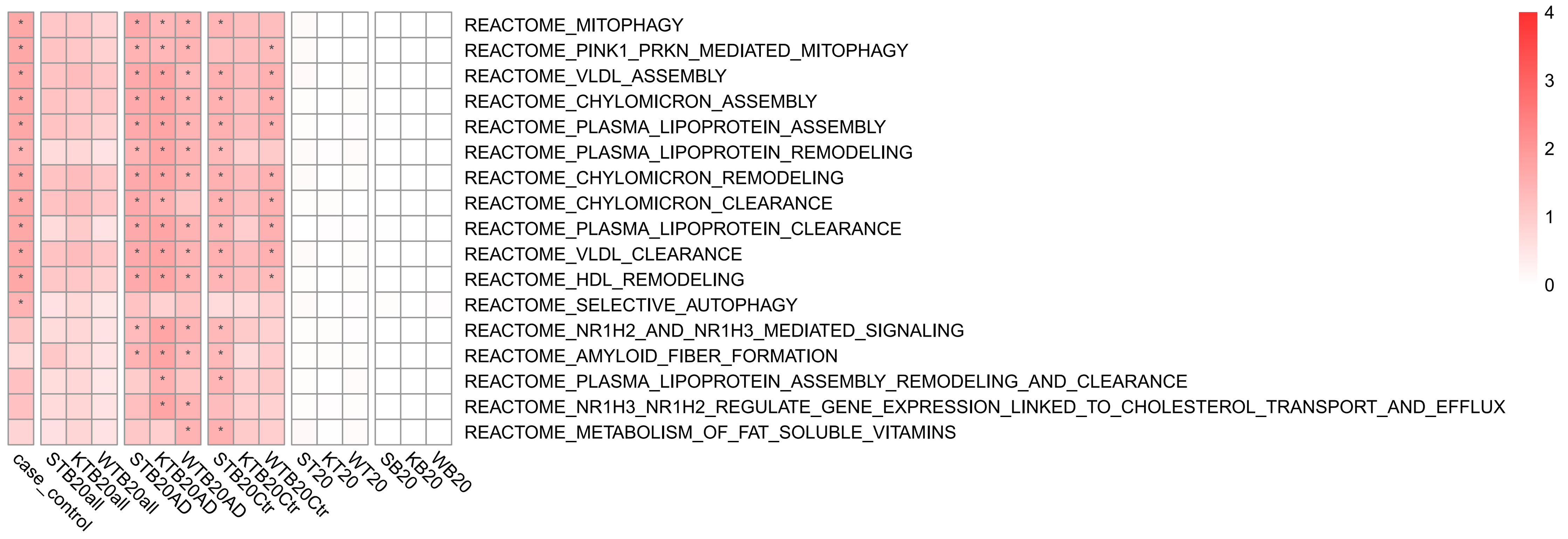


(C) Canonical Pathway PRSet identified in AD disc dataset Model 2

No significant GRPa in this condition.

(D) Canonical Pathway PRSet identified in AD rep dataset Model 2

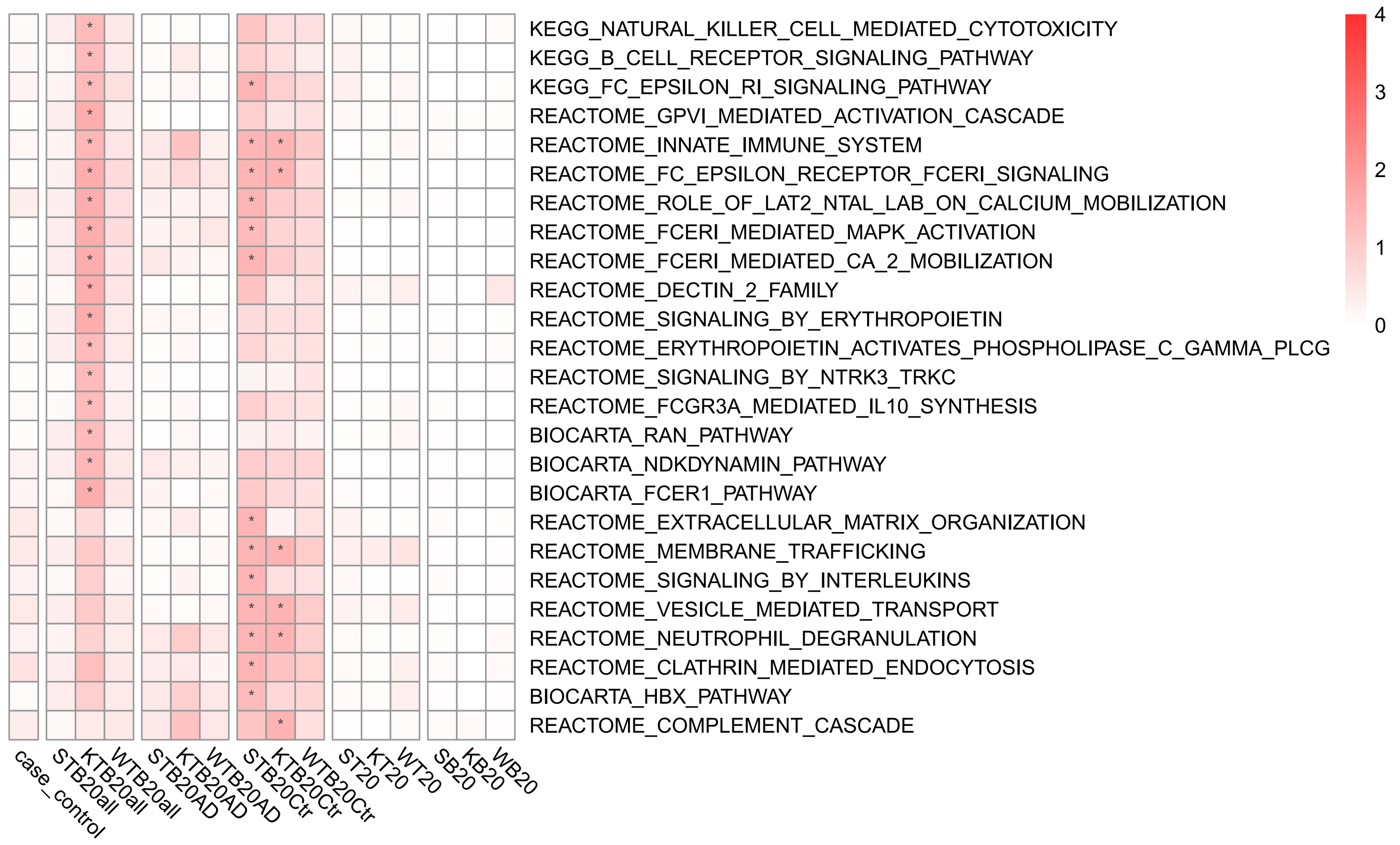


(E) Upset plot for overlapping signals between disc and rep datasets in Model 1.

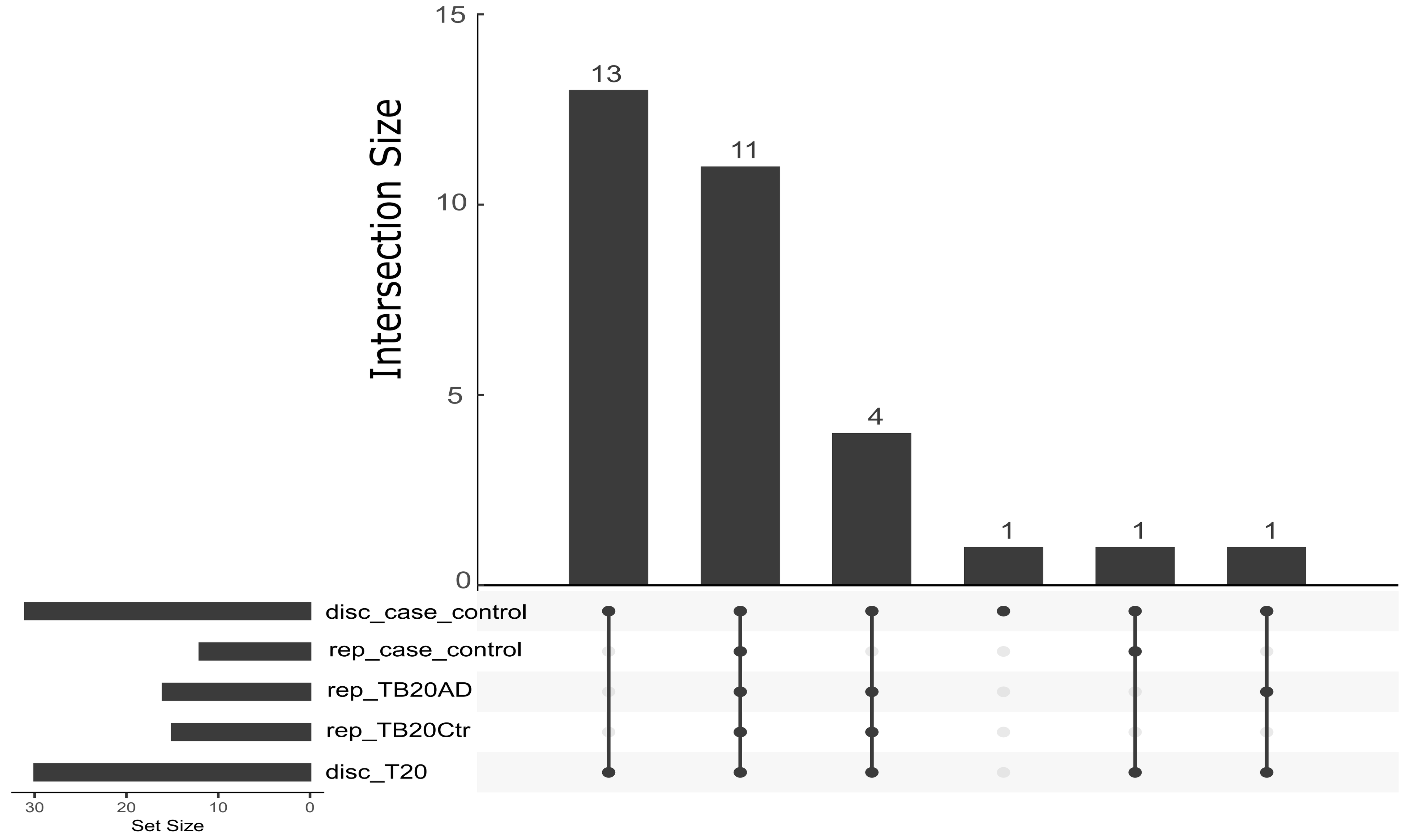
