## Supplemental Figure legends for "GRPa-PRS: A risk stratification method to identify genetically-regulated pathways in polygenic diseases"

**Supplementary figure legend**

**Fig. S1. MDS plot for individual ancestry along 1000 Genomes Project individuals.** (A), (B), (C), & (D) showed the MDS plot for four cohorts used in this study. We only selected individuals of European ancestry, and each plot uses the MDS threshold applied at the bottom left.

**Figure S2. PRS distribution for AD and SCZ cohorts.** The calculated PRS for both the discovery cohort and replication cohort is based on different Alzheimer’s Disease (AD) and Schizophrenia (SCZ) GWAS summary statistics. (A) PRS based on Schwartzentruber et al. GWAS for the AD discovery cohort; (B) PRS based on Kunkle et al. GWAS for the AD discovery cohort; (C) PRS based on Wightman et al. GWAS for the AD discovery cohort; (D) PRS based on Schwartzentruber et al. for the AD replication cohort; (E) PRS based on Kunkle et al. GWAS for the AD replication cohort; (F) PRS based on Wightman et al. GWAS for the AD replication cohort. (E) PRS based on Trubetskoy et al. GWAS for the SCZ SWE cohort; (F) PRS based on Trubetskoy et al. GWAS for the SCZ MGS cohort.

**Figure S3.** **Enrichment of GRPa-MAGMA results on AD brain function curation.** GRPas identified by GRPa-MAGMA are shown in the heatmap for (A) discovery cohort under Model 1 (full Model), (B) replication cohort under Model 1, (C) discovery cohort under Model 2 (no*-APOE* Model), and (D) replication cohort under Model 2 (no*-APOE* Model). * indicates the significant FDR < 0.05. Heatmap intensity indicates -log_10_(FDR). The x-axis shows the heatmap list of the subgroup comparison based on different GWAS summary statistics. S represents Schwartzentruber et al.; K represents Kunkle et al.; W represents Wightman et al..The overlapping signals between the discovery cohort and the replication cohort are shown in UpSet plot (E).

**Figure S4. Enrichment of GRPa-GSVA results on AD brain function curation.** GRPas identified by GRPa-GSVA are shown in the heatmap for (A) discovery cohort under Model 1 (full Model), (B) replication cohort under Model 1, (C) discovery cohort under Model 2 (no*-APOE* Model), and (D) replication cohort under Model 2 (no*-APOE* Model). * indicates the significant (p-value < 0.05 / # of gene set) GRPa identified in this condition. Heatmap intensity indicates -log_10_(p-value). The x-axis shows the heatmap list of the subgroup comparison based on different GWAS summary statistics. S represents Schwartzentruber et al.; K represents Kunkle et al.; W represents Wightman et al.. The overlapping signals between the discovery cohort and the replication cohort are shown in UpSet plot (E).

**Figure S5. Enrichment of GRPa-MAGMA results on AD canonical pathway curation.** GRPas identified by GRPa-MAGMA are shown in the heatmap for (A) discovery cohort under Model 1 (full Model), (B) replication cohort under Model 1, (C) discovery cohort under Model 2 (no*-APOE* Model), and (D) replication cohort under Model 2 (no*-APOE* Model). * indicates the significant FDR < 0.05. Heatmap intensity indicates -log_10_(FDR). The x-axis shows the heatmap list of the subgroup comparison based on different GWAS summary statistics. S represents Schwartzentruber et al.; K represents Kunkle et al.; W represents Wightman et al.. The overlap signals between the discovery cohort and the replication cohort are shown in UpSet plot (E).

**Figure S6. Enrichment of GRPa-MAGMA and GRPa-GSVA results on SCZ GO term curation.** GRPas identified by GRPa-MAGMA are shown in the heatmap for (A) SWE cohort and (B) MGS cohort. * indicates the significant GRPas FDR < 0.05. Heatmap intensity indicates -log_10_(FDR). GRPas identified by GRPa-GSVA are shown in the heatmap for (C) SWE cohort and (D) MGS cohort. * indicates the significant (p-value < 0.05 / # of gene set) GRPas identified in this condition. Heatmap intensity indicates -log_10_(p-value). The x-axis shows the heatmap list of the subgroup comparison based on different percentile threshold. (E), (F), & (G) show the semantic simillarity for significant terms from GRPa-MAGMA and GRPa-GSVA in BP, MF, and CC, respectively.

**Figure S7. Enrichment of GRPa-GSVA results on AD canonical pathway curation.** GRPas identified by GRPa-GSVA are shown in the heatmap for (A) discovery cohort under Model 1 (full Model), (B) replication cohort under Model 1 (full Model), (C) discovery cohort under Model 2 (no*-APOE* Model), and (D) replication cohort under Model 2 (no*-APOE* Model). * indicates the significant (p-value < 0.05 / # of gene set) GRPas identified in this condition. Heatmap intensity indicates -log_10_(p-value). The x-axis shows the heatmap list of the subgroup comparison based on different GWAS summary statistics. S represents Schwartzentruber et al.; K represents Kunkle et al.; W represents Wightman et al.. The overlapping signals between the discovery cohort and replication cohort are shown in UpSet plot (E).

**Figure S8. Enrichment of PRSet results on AD GO curation.** GRPas identified by PRSet are shown in the heatmap for (A) discovery cohort under Model 1 (full Model), (B) replication cohort under Model 1 (full Model), (C) discovery cohort under Model 2 (no*-APOE* Model) and (D) replication cohort under Model 2 (no*-APOE* Model). * indicates the significant GRPas FDR < 0.05. Heatmap intensity indicates -log_10_(FDR). The x-axis shows the heatmap list of the subgroup comparison based on different GWAS summary statistics. S represents Schwartzentruber et al.; K represents Kunkle et al.; W represents Wightman et al.. The overlapping signals between the discovery cohort and replication cohort are shown in UpSet plot (E).

**Figure S9. Enrichment of PRSet results on AD brain function curation.** GRPas identified by PRSet are shown in the heatmap for (A) discovery cohort under Model 1 (full Model), (B) replication cohort under Model 1 (full Model), (C) discovery cohort under Model 2 (no*-APOE* Model), and (D) replication cohort under Model 2 (no*-APOE* Model). * indicates the significant GRPas FDR < 0.05. Heatmap intensity indicates -log_10_(FDR). The x-axis shows the heatmap list of the subgroup comparison based on different GWAS summary statistics. S represents Schwartzentruber et al.; K represents Kunkle et al.; W represents Wightman et al.. The overlapping signals between the discovery cohort and replication cohort are shown in UpSet plot (E).

**Figure S10. Enrichment of PRSet results on AD canonical pathway curation.** GRPas identified by PRSet are shown in the heatmap for (A) discovery cohort under Model 1 (full Model), (B) replication cohort under Model 1 (full Model), (C) discovery cohort under Model 2 (no*-APOE* Model), and (D) replication cohort under Model 2 (no*-APOE* Model). * indicates the significant GRPas FDR < 0.05. Heatmap intensity indicates -log_10_(FDR). The x-axis shows the heatmap list of the subgroup comparison based on different GWAS summary statistics. S represents Schwartzentruber et al.; K represents Kunkle et al.; W represents Wightman et al.. The overlapping signals between the discovery cohort and replication cohort are shown in UpSet plot (E).

**Figure S11. UpSet plot for** **the overlapping signals GRPa-MAGMA, GRPa-GSVA, and PRSet in AD cohorts for AD brain function gene set curation.**

**Figure S12. UpSet plot for** **the overlapping signals GRPa-MAGMA, GRPa-GSVA, and PRSet in AD cohorts for canonical pathways curation.**

**Figure S13. Heatmap for top 20 significant MultiXcan gene for each strata comparison in GO term curation for AD and SCZ.** (A) AD discovery cohort, (B) AD replication cohort, (C) SCZ SWE cohort, and (D) SCZ MGS cohort. The intensity is -log_10_(FDR) of gene in each strata comprison. * indicates the signficant MultiXcan gene FDR < 0.05.

**Figure S14.** **Heatmap for top 5 GWAS p-value genes from each signifcant GRPa across all significant GO terms** (A) significant GO BP terms from GRPa-GSVA in AD disc and rep cohorts, (B) significant GO MF terms from GRPa-GSVA in AD disc and rep cohorts, (C) significant GO CC terms from GRPa-GSVA in AD disc and rep cohorts, and (D) significant GO terms from GRPa-MAGMA and GRPa-GSVA in SCZ SWE and MGS cohorts. The intensity is -log_10_(p-value), where gene p-value is calculated from Wightman et al. GWAS using MAGMA.

**Figure S15.** **Orthgonoal test for individual** **PRS correlation and gene set** **GSVA score in subgroups.**  The correlation between sPRS and gene set (A) Microglia Cell death & apoptosis, and (C) BP myelin maintenance activity. (B) and (D) show the distribution differences of GSVA score within group 1 versus group 2 comparisons among subgroups STB20all, STB20AD, STB20Ctr, and ST20.

**Figure S16.** **Effect size and power analysis.** (A) Boxplot of absolute effect sizes for different percentile subgroups (10, 15, and 20) in AD discovery TBall stratum, covering all GO terms across 13 brain tissues and three parallel PRS subgroups (S,K,W). S represents Schwartzentruber et al.; K represents Kunkle et al.; W represents Wightman et al.. (B) Boxplot of absolute effect sizes for different percentile subgroups (10, 15, and 20) in SCZ TBall stratum, covering all GO terms across the 13 brain tissues and two cohorts (SWE and MGS). (C) Power analysis for assessing the various effect sizes and the sample sizes required at the alpha of 0.05.

**Figure S17.** **Enrichment of GRPa-MAGMA results on GO curation for APOE allele stratified case control comparison.**  GRPas identified by GRPa-MAGMA for stratified case-control comparison using APOE genotype of individuals (22,23,33,34,44) are shown in the heatmap for (A) discovery cohort in GO terms, (B) replication cohort in GO terms, (C) discovery cohort in canonical pathways, (D) replication cohort in canonical pathways, (E) discovery cohort in AD brain function gene sets, and (F) replication cohort in AD brain function gene sets. * indicates the significant FDR < 0.05. Heatmap intensity indicates -log_10_(FDR). The x-axis shows the heatmap list of the case-control strata comparison stratified by individual APOE genotype.

**Figure S18.** **Enrichment analysis of GRPa-MAGMA results with PRS value adjustion for three gene set curations.** GRPas identified by GRPa-MAGMA with PRS value adjusted for (A)-(E). * indicates the significant FDR < 0.05. Heatmap intensity indicates -log_10_(FDR). The x-axis shows the heatmap list of the subgroup comparison based on different GWAS summary statistics. S represents Schwartzentruber et al.; K represents Kunkle et al.; W represents Wightman et al..
